# Topical Steroid Withdrawal is a Targetable Excess of Mitochondrial NAD+

**DOI:** 10.1101/2024.04.17.24305846

**Authors:** Nadia Shobnam, Sarini Saksena, Grace Ratley, Manoj Yadav, Prem Prashant Chaudhary, Ashleigh A Sun, Katherine N Howe, Manasi Gadkari, Luis M Franco, Sundar Ganesan, Katelyn J McCann, Amy P Hsu, Kishore Kanakabandi, Stacy Ricklefs, Justin Lack, Weiming Yu, Morgan Similuk, Magdalena A Walkiewicz, NIAID Centralized Sequencing Program, Donna D Gardner, Kelly Barta, Kathryn Tullos, Ian A Myles

## Abstract

**Background:** Topical corticosteroids (TCS) are first-line therapies for numerous skin conditions. Topical Steroid Withdrawal (TSW) is a controversial diagnosis advocated by patients with prolonged TCS exposure who report severe systemic reactions upon treatment cessation. However, to date there have been no systematic clinical or mechanistic studies to distinguish TSW from other eczematous disorders.

**Methods:** A re-analysis of a previous survey with eczematous skin disease was performed to evaluate potential TSW distinguishing symptoms. We subsequently conducted a pilot study of 16 patients fitting the proposed diagnostic criteria. We then performed: tissue metabolomics, transcriptomics, and immunostaining on skin biopsies; serum metabolomics and cytokine assessments; shotgun metagenomics on microbiome skin swabs; genome sequencing; followed by functional, mechanistic studies using human skin cell lines and mice.

**Results:** Clinically distinct TSW symptoms included burning, flushing, and thermodysregulation. Metabolomics and transcriptomics both implicated elevated NAD+ oxidation stemming from increased expression of mitochondrial complex I and conversion of tryptophan into kynurenine metabolites. These abnormalities were induced by glucocorticoid exposure both *in vitro* and in a cohort of healthy controls (N=19) exposed to TCS. Targeting complex I via either metformin or the herbal compound berberine improved outcomes in both cell culture and in an open-label case series for patients with TSW.

**Conclusion:** Taken together, our results suggest that TSW has a distinct dermatopathology. While future studies are needed to validate these results in larger cohorts, this work provides the first mechanistic evaluation into TSW pathology, and offers insights into clinical identification, pharmacogenomic candidates, and directed therapeutic strategies.

## Introduction

Topical steroids are first-line therapies for atopic dermatitis (AD) and other inflammatory dermatological conditions^1^. Although more accurately described as glucocorticoids, topical corticosteroids (TCS) is the more common clinical term. In recent decades, there has been a concerning rise in reports of severe systemic adverse reactions due to long-term use and abrupt cessation of TCS (commonly referred to as Topical Steroid Withdrawal; TSW)^2^. Although no formal diagnostic criteria for TSW exist, reports of patients experiencing TSW are very common online^3^. Initial reports concluded that TSW was limited to exposed areas for patients who applied high-potency TCS to the face or genitals^4^. However, subsequent case series have better identified the ways in which TSW may be different from AD, contact dermatitis, or other dermatoses^2,5–7^, including disease manifestations on regions of the body where TCS were never applied. Although resolution may take months to years, improvement is seen through avoidance of TCS therapy, further suggesting that TSW is a distinct clinical entity rather than a flare of the underlying dermatoses^8^.

Although some have labeled the patient concerns as “steroid phobia”^9^, there have been no systematic clinical or mechanistic studies to either refute or support TSW as molecularly distinct from other dermatopathies. Herein, we report a re-analysis of a previous survey of 1,889 adults with eczematous skin disorders^7^ to clinically distinguish those that self-diagnosed with TSW from those that denied having TSW. We then report the results of a pilot study of 16 patients with symptoms consistent with TSW. Using multi-omics analysis of serum and skin samples we identified increased vitamin B3 (nicotinic acid; NAD+) oxidation stemming from overexpression of known NAD+ producers^10,11^ such as mitochondrial complex I. Open-label treatment with anti-complex I medications (metformin or the “herbal metformin” berberine) led to notable improvements in symptoms. Our work represents the first investigation into the pathogenesis of TSW resulting in a promising targeted therapy.

## Results

### Patients with TSW have symptoms distinct from atopic dermatitis

Reanalysis of a previously published survey of patients with eczematous skin disease^7^ identified worsening skin symptoms despite TCS use as most predictive of self-diagnosed TSW compared to those with eczematous skin disease who denied having TSW (Fig. 1A; Fig. S1A). Worsening signs may include reduced TCS effectiveness, a need for higher potency, spreading rash despite treatment, or new-onset symptoms of temperature dysregulation and full body redness/burning (Fig. 1A). Peak itch intensity and sleep disturbance were significantly higher in those affirming TSW (Fig. 1B). However, there were no consistent, significant differences in body site for either severity (Fig. S1B) or application site (Fig. 1A). The dryness and oozing scores typically collected for the Patient Oriented SCORing Atopic Dermatitis (PO-SCORAD^12^) severity metric were inversely associated with TSW (Fig. 1A), however a greater degree of redness, scratching, swelling, and thickening were seen in TSW (Fig. S1C). Skin specific symptoms with a greater than 80% prevalence that are distinct from AD were deemed as major criteria while the remainder were utilized as minor criteria (Fig. 1C). Twenty-five percent of patients reported all related symptoms, while the combination of ≥1 major and ≥3 minor offered >90% sensitivity for capturing those reporting TSW in the survey (Fig. 1C). Nearly 25% of patients reported symptoms lasting longer than 3 years after discontinuation of TCS (Fig. 1D).

**Figure 1:**
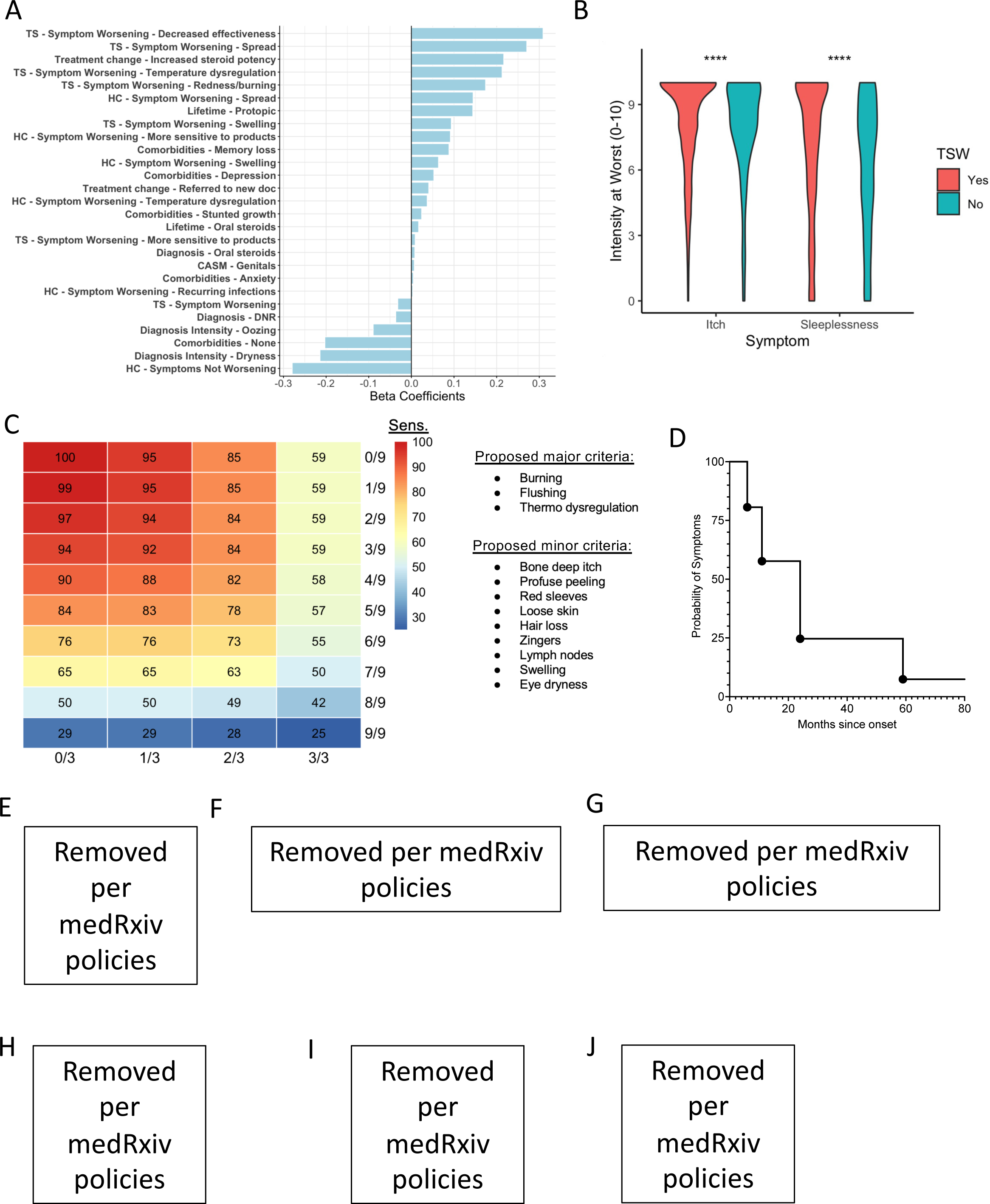
Topical Steroid Withdrawal (TSW) is distinguishable from other eczematous skin diseases. (A) Lasso regression model of previously reported survey results from 1,889 individuals with eczematous skin disease^7^ establishing the responses that are most predictive of self-identified TSW. Results separated by over-the-counter hydrocortisone (HC) versus prescription topical steroids (TS). (B) Intensity of itch and sleep disruption for the respondents affirming TSW versus those denying such diagnosis. (C) Proposed diagnostic criteria derivation evaluating the sensitivity (Sens.) for number of reported Major and Minor criteria extracted from survey from respondents reporting suffering from TSW (N=1,486). (D) Time to symptom of TSW symptoms as reported by respondents (N=1,486). (E-I) Representative images of TSW associated symptoms; although images provided with explicit permission and consent, medRxiv does not allow images of body parts and thus these images have been removed. The final publication will display these images.

Although many reports imply that TSW is limited to the application of high-potency glucocorticoids to the face or genitals^4,13^, we enrolled a cohort (N = 16) who each experienced full-body disease, including areas that had never been directly treated with TCS (Fig. S1D; Table S1). Patients described full body redness, flushing, and anhidrosis (Fig. 1E; redacted per medRxiv policies) occurring 4-6 weeks after discontinuation of TCS, which: tended to spare the nose (Fig. 1F; termed “headlamp sign”^14^; redacted per medRxiv policies), palms, and soles (Fig. 1F; redacted per medRxiv policies); and often formed “red sleeves” on the arms (Fig. 1G; redacted per medRxiv policies). Additional symptoms included: subjective temperature changes; stabbing neuropathic pain (termed “zingers” by patients; Fig. S1D); loose skin over the flexor surfaces (“elephant wrinkles”; Fig. 1H; redacted per medRxiv policies); hair loss (Fig. 1I; redacted per medRxiv policies); and substantial skin shedding (Fig. 1J-redacted per medRxiv policies; Fig. S1D).

### Serum metabolomics suggest systemic lipid dysregulation

Serum metabolomics for patients with TSW was compared to healthy volunteers (HV; N = 11), and patients with AD who did not report symptoms of TSW (N = 10). Although matched for sex and ethnicity, the TSW cohort was slightly older (mean 37.1 years) than AD (26 years) and HV (25 years; Table S1; redacted per medRxiv policies). Significant differences were seen under global similarity analysis but not for any specific identifiable metabolites (Fig. S2A-D). However, collating the results into MetaboAnalyst pathway analysis^15^ suggested patients with TSW had significant deficiencies in the sphingolipid and urea cycle amino acid pathways (Fig. S2E) compared to both HV and AD. Patients with TSW had significant overabundance of metabolites in several pathways, including glycerophospholipids and C21-steroid hormones including cortisol (Fig. S2F; Fig. S3).

### Markers of dysbiosis and inflammation in TSW were similar to those previously described in atopic dermatitis

Evaluation of the microbiome signatures in TSW revealed significant differences from HV in bacterial speciation at the genus level (Fig. 2A-B; Fig. S4A). Similar to AD^16^, there was a predominance of Staphylococcal species, particularly *S. aureus*, burden (Fig. 2C) and reduced alpha bacterial diversity (Fig. 2D). Less robust differences with respect to HV were seen in fungal speciation, with results again similar to those described for AD (Fig. 2E; Fig. S4B)^17^. Virome signatures demonstrated an increased burden of viruses associated with fungal and bacterial taxa (Fig. S4C) while functional metagenomics revealed global differences (Fig. 2F) that were dominated by transcripts associated with *Staphylococcus* infection (Fig. S5A-B).

**Figure 2:**
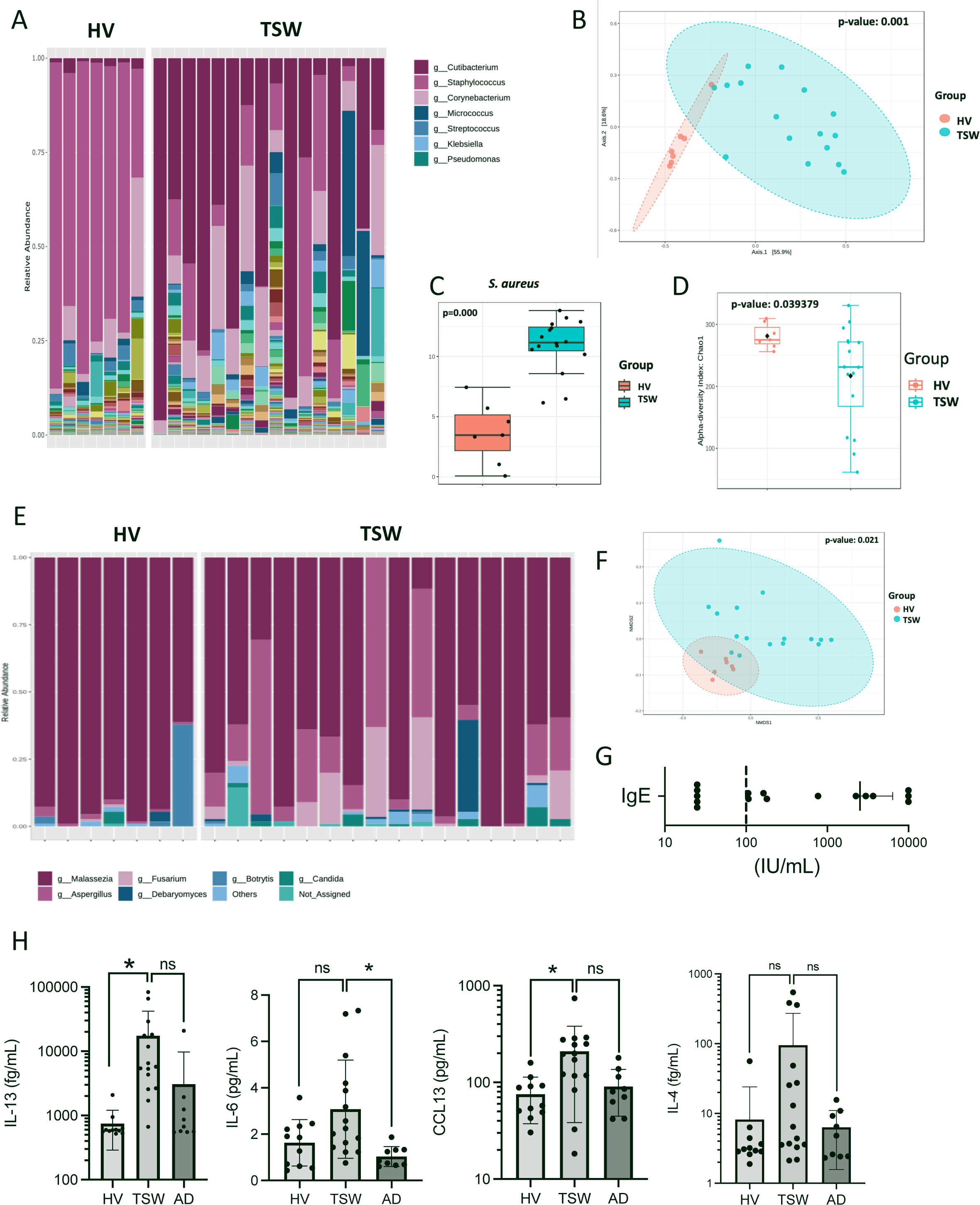
Microbiome and Th2 cytokine signatures in TSW differ from healthy controls. Shotgun metagenomic sequencing was performed on skin swabs from patients with TSW (N=16) versus healthy volunteers (N=7). (A) Speciation at genre level with most abundant taxa indicated (full list in Fig. S4A). (B) Similarity plot as measured by NMDS (non-metric multidimensional scaling) for genre-level bacterial analysis. (C) Normalized counts for *Staphylococcus aureus*. (D) Chao1 alpha-diversity index. (E-F) Speciation and NMDS for fungal genre. (G) IgE levels (mean ± SD) for TSW cohort (dotted line indicates upper limit of normal for our hospital). (H) Serum cytokine levels for HV, TSW and patients with atopic dermatitis without history of TSW (AD) for interleukin (IL-) 13, IL-6, CCL13, and IL-4. Significance adjusted for three-way comparison, but not the full number of cytokines analyzed. * = p value <0.05, ns = not significant, other p values indicated.

Like AD^18^, preliminary characterization of cell infiltrates were primarily CD4+ and CD8+ T-cells, as well as CD68+ monocytes (Fig. S5C); these immune cells were detectable in thin sections of skin biopsies for patients with TSW but required 3-dimensional volume analysis to detect in HV samples (Fig. S5D; Movie S1). For 11 of the patients in the cohort, IgE levels were elevated (Fig. 2G). Consistent with reported utility of dupilumab in TSW^19^, serum cytokine analysis indicated significantly increased levels of interleukin (IL-) 4 and IL-13, as well as proinflammatory IL-6 and CCL13 (Fig. 2H; Table S2).

### Metabolomics implicates vitamin B3 metabolism

Skin metabolomics was performed using MALDI-imaging mass spectrometry^20^, which identifies metabolites in space (example shown in Fig. 3A). Ranking the abundance of metabolites that could be identified using m/z and collisional cross section (CCS) revealed distinctions between TSW and HV (Fig. 3B). Using MetaboAnalyst^15^ to contrast TSW with both HV and AD identified a significant increase in vitamin B3 (nicotinate and nicotinamide) metabolism (Fig. 3C). Several pathways were downregulated compared to the HV and AD cohorts, most prominently tryptophan metabolism (Fig. 3D). Of note, nicotinic acid is a derivative of tryptophan through a process that generates the known neuropathologic, and anticholinergic intermediate kynurenine via the enzymes indoleamine 2,3-dixygenase (IDO) and tryptophan 2,3-dioxygenase (TDO)^10^. Consistent with the known interaction between NAD+ and the itch regulating transient receptor potential (TRP) family ion channels^21,22^, staining a subset of patients and controls for expression of TRPA1 revealed enhanced dermal, but not total, expression (Fig. S5E-F).

**Figure 3:**
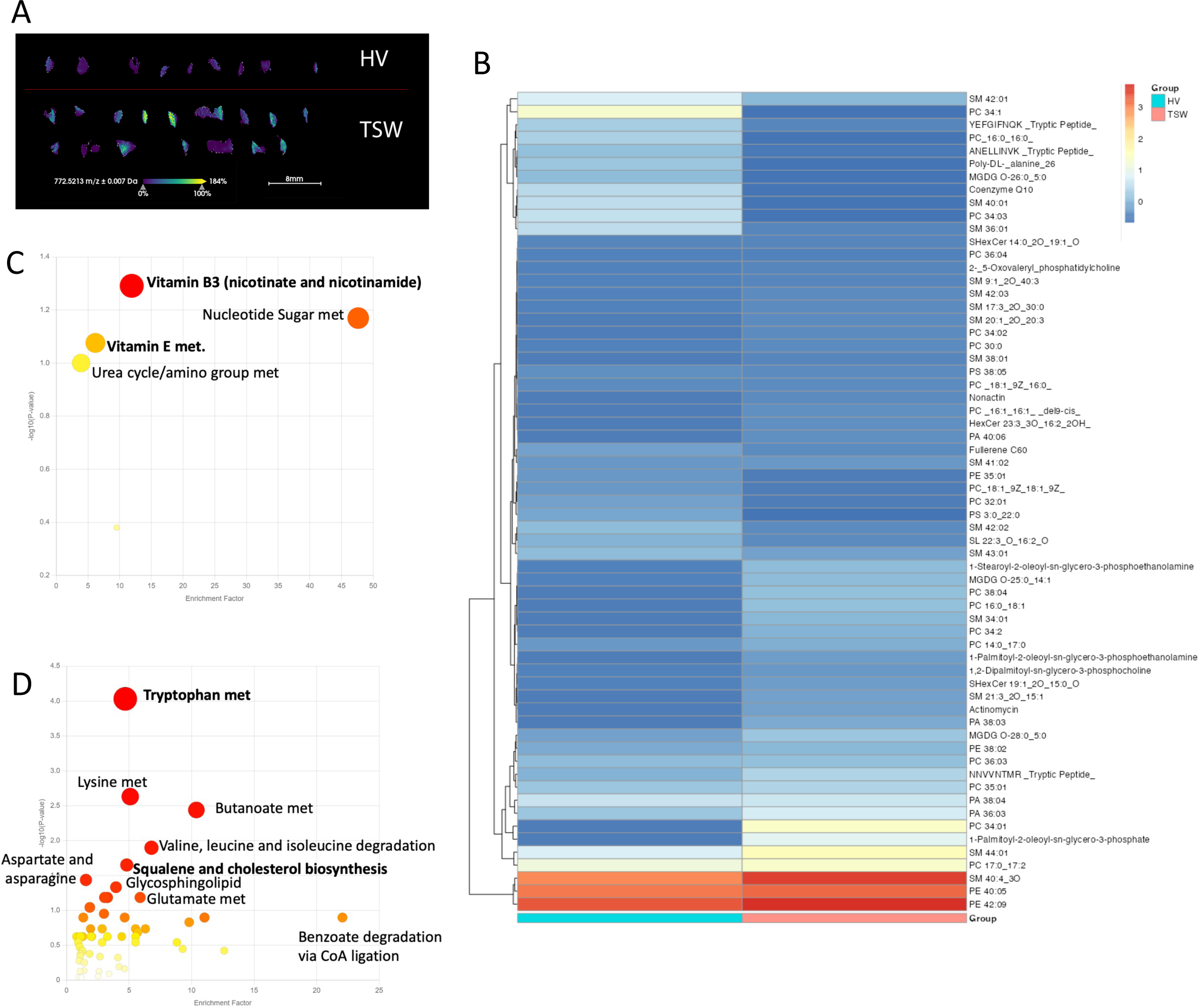
Skin from patients with TSW demonstrates disruptions in Vitamin B3 and Tryptophan metabolism. Skin biopsy samples from patients with topical steroid withdrawal (TSW; N=16), Atopic Dermatitis (AD; N=10), and healthy volunteers (HV; N=11) underwent MALDI-imaging mass spec. (A) Representative image for distribution of metabolite at 772.5213 m/z. for HV and TSW (B) Intensity values for metabolites identifiable through combination of m/z and collisional cross-section for HV and TSW. (C-D) MetaboAnalyst pathway analysis for metabolites ranked by how predictive of TSW they were compared with both HV and AD cohorts (C) and metabolites ranked by ROC value (receiver operating characteristic) for the HV and AD cohorts compared to TSW (D). Bolding indicates pathways not identified as distinctive for AD relative to HV.

### RNAseq suggests mitochondrial dysfunctionis present in TSW

Biopsies subjected to RNAseq identified cell cycle and neurodegeneration as the most distinguishing pathways when comparing TSW to HV (Fig. S6A). The neurodegeneration pathway includes *Wnt* signaling (Fig. 4A) which has been linked to AD previously and would be expected to produce increased cell proliferation^23^. Several mitochondrial complexes were also upregulated in patients with TSW, particularly complex I (Fig. 4B) which produces NAD+ as a byproduct. Several other aspects of nicotinate metabolism were upregulated in TSW (Fig. S6B). Opposing expression of *IDO2* and *TDO2* was seen between TSW and controls (Fig. 4C; full image in Fig. S6C). STRING analysis^24^ demonstrated a strong signal for histone modifying and DNA processing pathways but were most strongly linked to proton motive force in ATP synthesis (Fig. S6D). Activation of inflammatory pathways known to respond to upregulations in the oxidative stress pathways (Fig. S6A) was evident for MAP kinase (Fig. S7A), JAK-STAT (Fig. S7B), NFκB (Fig. S8A), and T-cell receptor signaling (Fig. S8B).

**Figure 4:**
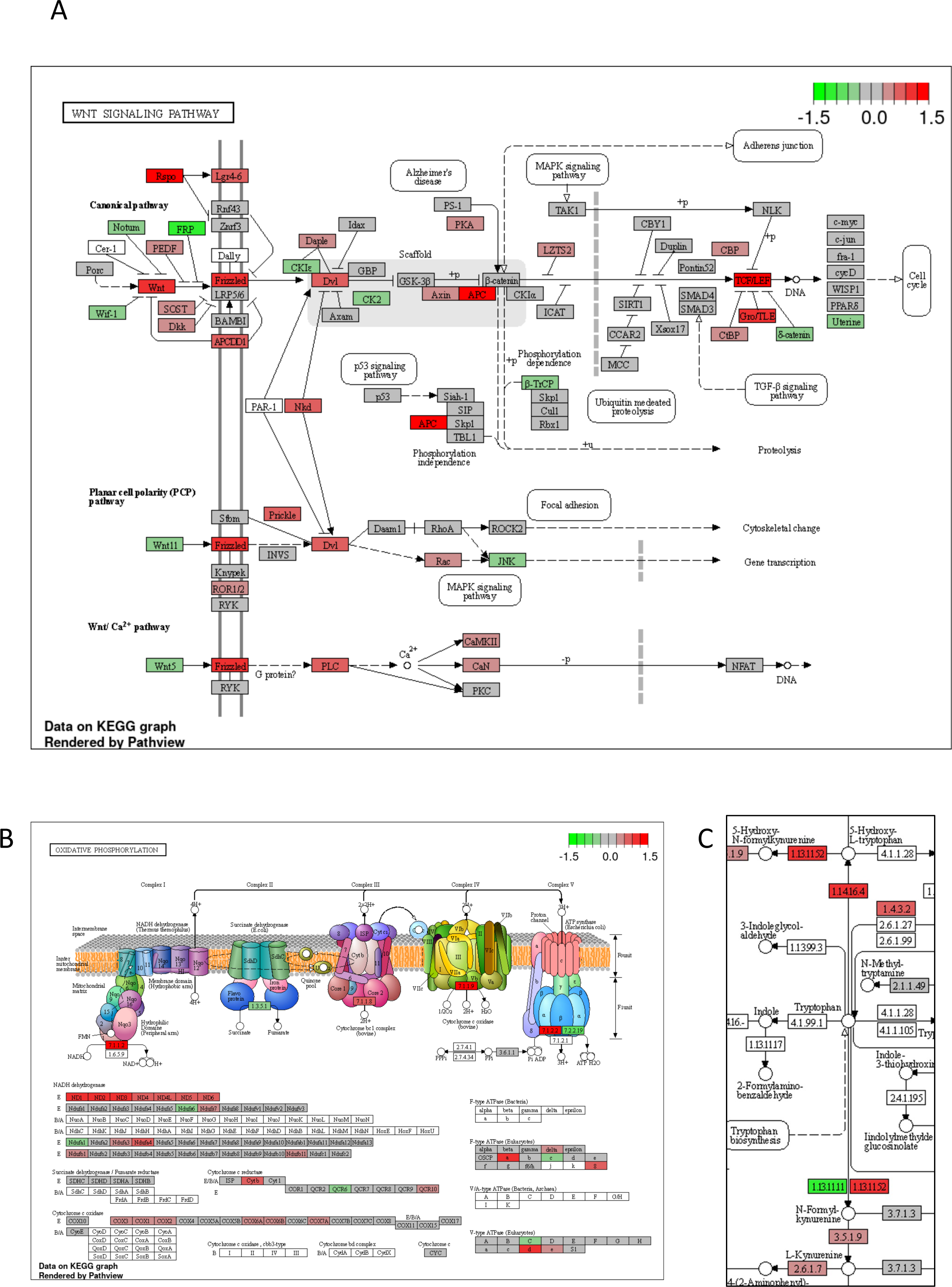
Skin from patients with TSW demonstrates disruptions in Niacin related metabolic pathways. Skin biopsy samples from patients with topical steroid withdrawal (TSW; N=16) and healthy volunteers (HV; N=11) underwent RNAseq analysis. (A-C) KEGG pathway analysis for *Wnt* signaling (A), Oxidative phosphorylation (B), and Nicotinate and Nicotinamide metabolism (C; full image in Fig. S6B). Superimposed log_10_ fold change, green indicates product is more abundant in HV, red indicates product is more abundant in patients with TSW.

### Nicotinic acid and clobetasol exposure impact stem cell metabolism

Although niacin and nicotinic acid are known to induce skin flushing^25^, most research into the skin effects of niacin has focused on dietary intake. Topical application of nicotinic acid to mouse ears yielded a dose-dependent increase in swelling (Fig. 5A), but with redness that was below the mouse hairline (Fig. 5B). This finding is distinct from the standard murine models of AD such as MC903, which induce dermatitis on the entire ear (Fig. S9A). Combining the prior reports of nasal, palmar, and plantar sparing, the protracted resolution^14^, and murine disease limited to areas with hair (Fig. 5B) suggested a possible role for stem cells in the pathology of TSW given that these cells reside at the base of each hair follicle and serve as progenitors for replenishing other skin cells^26^.

**Figure 5:**
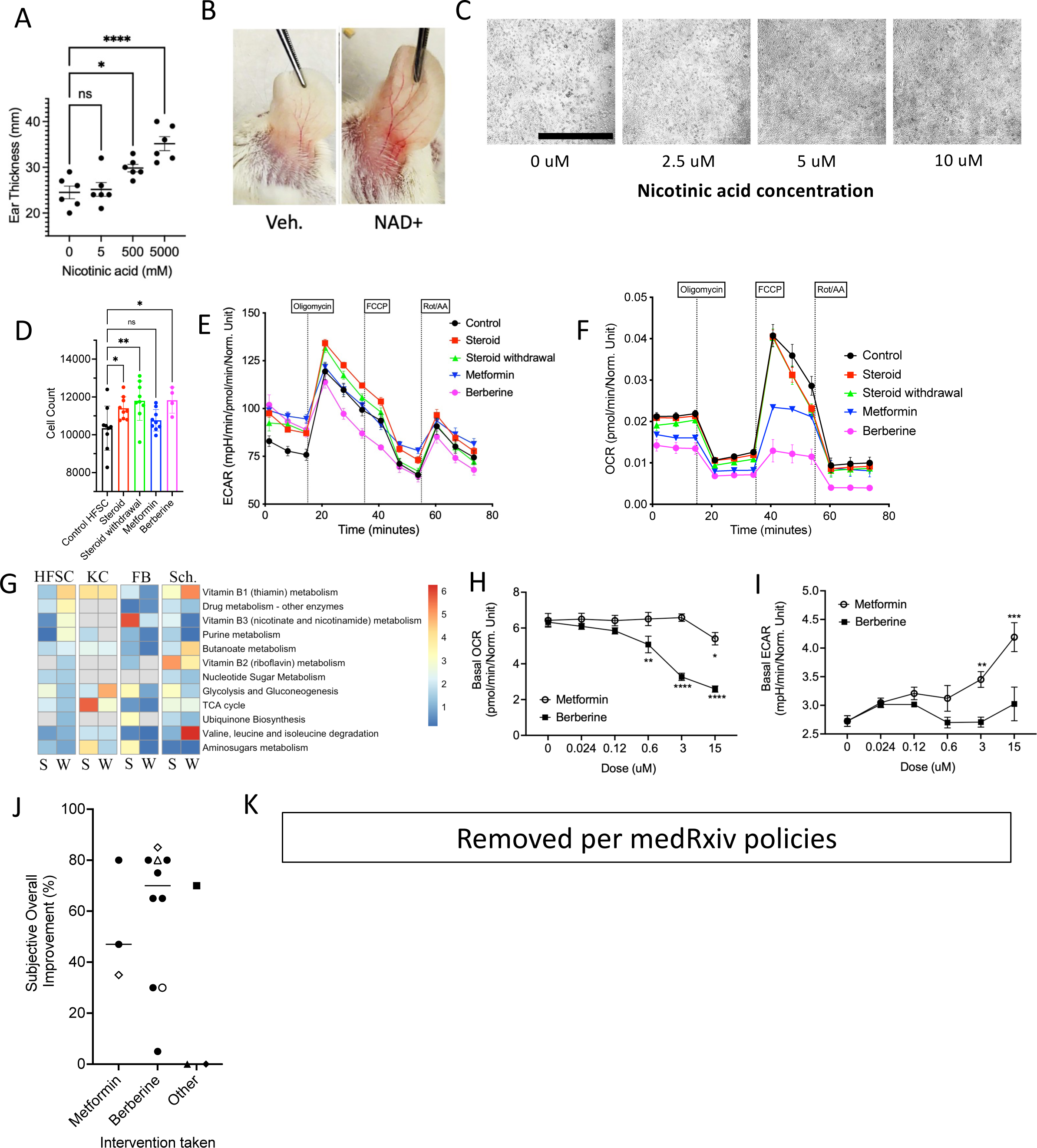
Steroid withdrawal induces mitochondrial associated niacin overproduction in human skin cells. (A-B) Mice were treated for 8 days with daily topical application of nicotinic acid. Resultant ear thickness (A) and representative image of redness below shaved hairline (B) are shown. (C) Human follicular stem cells (HFSC) exposed to increasing doses of nicotinic acid, scale bars indicate 2mm. (D) Total cell count for HFSC incubated in clobetasol for 48 hours (Steroid), clobetasol for 24 hours followed by normal media for 24 hours (Steroid withdrawal), or under steroid conditions with 10uM for metformin or berberine. Dots indicate replicate wells. (E-F) HFSC under mitochondrial stress test for Seahorse assay; indicating basal extracellular acidification rate (ECAR; a measurement of glycolysis) and basal oxygen consumption rate (OCR; a proxy for oxidative phosphorylation) for cells incubated as in panel D. (G) MetaboAnalyst pathway derive index of pathway significance for HFSC, keratinocytes (KC), fibroblasts (FB), or Schwann cells (Sch.) under steroid (S) and steroid withdrawal (W) conditions as measured by imaging mass spec. (H) Basal OCR and ECAR for HFSC incubated with metformin or berberine (but no glucocorticoids) for 24 hours before Seahorse analysis. (J) Response from patients for global assessment of overall improvement after 3-5 months of treatment with indicated intervention. Open diamond indicates patient who started on metformin for 2 months, then switched to berberine for 2 months. Open circle indicates patient that discontinued berberine after 4 weeks when she discovered she was pregnant. Open triangle indicates a patient who used both berberine and ‘activated plasma’. Closed square indicate patient with a multitude of various interventions (See Table S3 for more details). One participant was loss to follow up. (K) Representative images of improvement in patients who used berberine; although images provided with explicit permission and consent, medRxiv does not allow images of body parts and thus these images have been removed. The final publication will display these images. Data represent 3 independent experiments (A-I) and are indicated by mean ± SEM.

Similar to prior reports^27^, incubation of human follicular stem cells (HFSC) with nicotinic acid led to enhanced proliferation (Fig. 5C). Exposure to the commonly prescribed, high-potency glucocorticoid clobetasol also significantly increased HFSC proliferation (Fig. 5D). Functional metabolic analysis of HFSC indicated clobetasol exposure increased maximal extracellular acidification rate (ECAR; a measurement of glycolysis; Fig. 5E, Fig. S9B) but not resting oxygen consumption rate (OCR; a proxy for oxidative phosphorylation; Fig. 5F, Fig. S9C). Incubation of HFSC with clobetasol did not impact vitamin B3 metabolism, however, removing steroids from the media after clobetasol exposure impacted vitamin B3 and other metabolic pathways including the TCA cycle (Fig. 5G; Fig. S9D). Differences between continuous glucocorticoid exposure and steroid withdrawal were also evident for human primary keratinocytes, fibroblasts, and Schwann nerve cells (Fig. 5G; Fig. S9E-G).

These results suggested that TSW pathology may represent an overactivation of complex I driving up NAD+ oxidation, either through increased complex I expression or increased NADH availability via tryptophan metabolism (Fig. S10A). Metformin is a known blocker of complex I^11^, has pre-clinical efficacy in models of AD^28^, and reduces the conversion of tryptophan to kynurenine^29^. Similarly, berberine is the main ingredient in the traditional Chinese medicine, Wu Mei Wan, previously shown to improve TSW^30^; berberine is also a known complex I inhibitor with similar uses as metformin^31^. Metformin treatment of HFSC prevented hyperproliferation (Fig. 5D) as well as the alterations in basal OCR and ECAR (Fig. 5E-F, Fig. 5H-I, Fig. S9B-C). Berberine did not alter proliferation (Fig. 5D) or basal ECAR (Fig. 5I, Fig. S9B) but was a more potent inhibitor of OCR (Fig. 5F, Fig. 5H, Fig. S9C). Diluting berberine to as low as 0.6uM had inhibitory activity without a strong coloration (Fig. 5H, Fig. S10B).

### Topical glucocorticoids alter mitochondrial transcription pathways

Nineteen healthy controls aged 18-64 were treated with the topical glucocorticoid methylprednisolone aceponate. Skin biopsies were taken prior to exposure, then again 4 hours after topical application. RNAseq identified numerous significant differences (Fig. S11A) concentrated in the keratinocyte differentiation pathways (Fig. S11B). Mitochondrial complexes, especially the complex I, NAD+ producing enzyme, ubiquinone reductase were significantly upregulated by topical glucocorticoid exposure (Fig. S11C-D). Impacts on *Wnt* and tryptophan metabolism were evident, but less robust than for TSW (Fig. S12A-B).

### Complex I Targeting Therapies Improve TSW Symptoms

Patients in the cohort were informed about the potential utility of metformin and berberine and provided the analysis by the commercial outlet ConsumerLabs identifying that Natural Factors WellBetX and Solaray supplement brands contained the indicated amount of berberine without harmful contaminants. Three patients elected to take metformin through their primary care providers, 9 elected to take berberine purchased online, 2 made no changes in treatment, while 1 opted for alternate treatments (Table S1). No serious complications were reported (Table S1).

After 3-5 months of use, subjective overall improvement between 5% and 80% was reported for both interventions (Fig. 5J), with non-significantly greater improvement reported for those on berberine. Patients initially noted their skin becoming “softer” and/or “smoother” followed by an “inside out” resolution of itch in which the bone-deep itch resolved before the superficial itch. Rashes improved in both groups (Fig. 5K; redacted per medRxiv policies). Patients still reported flares related to known triggers such as infection, stress, or temperature change; however, these flares were described as of lower severity or shorter duration.

### DNA variants with differentially expressed transcripts in TSW included PAX6 and FMO2

Genome sequencing only identified the highly variable *TTN* as the gene with variants of uncertain significance (VUS) across the cohort (Fig. S13A-B). Expanded analysis of the genome identified 206 genes with DNA variants present in at least 12 of the 16 participants with combined annotation dependent depletion (CADD) scores greater than 20 (a predictor of how deleterious to protein function a variant might be; Fig. S13C). STRING analysis of these variants suggested the corresponding genes were enriched for involvement in cell adhesion, neuron development, and mitochondrial depletion (Fig. S13D-F). Comparing the variant list with the RNAseq results indicated that genes which contained DNA variants and differentially expressed transcripts were led by the known modulator of NADPH metabolism *FMO2*^32^ and *PAX6*, which may mediate AD-relevant pathways in epithelial-to-mesenchymal transition^33,34^ (Fig. S13G-I). Sequence analysis of the mitochondrial genomes did not identify any common non-synonymous variants within our cohort (Table S3).

## Discussion

Our results indicate that TSW is a distinct, iatrogenic dermatopathy deserving of further investigation. In this study, we propose diagnostic criteria to separate TSW from other eczematous skin disorders, provide early insights into cellular pathogenesis, identify candidate DNA variants, and describe the results from an open-label case series of two molecularly targeted therapies. Although the severity of skin disease was greater in our TSW cohort than their AD comparators, our findings are distinct from the numerous reports of lipid-centric defects seen in severe AD^35^.

Our working hypothesis of TCS induced complex I hyperactivity in follicular stem cells is supported by the established effects of glucocorticoids on mitochondria^36^ and the clinical data detailing the distribution and duration of TSW symptoms^2,5–7^. In some individuals, prolonged use may result in a form of endogenous chemical irritation due to overproduction of oxidized NADH (NAD+). The enhanced NADH production generated by tryptophan degradation results in an increase in neuro-toxic kynurenine metabolites via the corticosteroid-responsive enzymes IDO and TDO^10^. We hypothesize that epigenetic changes may underly the protracted nature of TSW recovery, however cell specific analysis will be needed to further elucidate our findings.

A hypothesis predicated on NAD+ is consistent with each of the proposed phases of TSW^7^: the initial rise in NAD+ combined with the resultant anticholinergic effects of kynurenic mediators may lead to the early signs of flushing, anhidrosis, palpitations, neuralgia, and vision changes^37^. Subsequent Wnt overactivity, mitochondrial inflammatory reactive oxygen species, and TRP receptor activation would be expected to induce desquamation, pruritus, skin proliferation, changes in temperature sensation, and may make those with TSW more susceptible to effects of chemical triggers associated with pruritic skin diseases^38^.

Our work suggests clinical insights for practitioners treating patients experiencing worsening eczematous skin disease despite >4 months of TCS treatment. For example, the prior claims of disease limited to the face or genitals^4^ are not supported by our data and should not guide diagnosis. Instead, patients with the identified TSW predictive symptoms should be evaluated for the diagnosis and considered for treatment using mitochondrial complex I inhibitors; however, expanded formal trials will be needed to confirm the safety and efficacy of these treatments. Although topical berberine was not attempted in this study, the efficacious concentrations *in vitro* are akin to dissolving one 500 mg capsule in a liter of water and then adding ¼ cup of the resultant solution to a standard 60-gallon bathtub as is done with bleach baths in AD^39^. Exposure to heat may also be a speculative intervention given higher temperatures preferentially inhibit mitochondrial complex I^40^.

Our work is limited in several ways. First, querying TSW-specific symptoms from eczematous patients who deny having TSW will be required to validate the diagnostic criteria through calculating specificity and accuracy. The small pilot enrollment and open label case series may not fully capture either the pathology or treatment response for a disorder with a reported population of the scale of TSW. Our global subjective assessment of TSW improvement should be replaced by prospective severity indices specific to TSW which incorporate scores for thermodysregulation, burning, and flushing. Additionally, larger studies will be needed to test whether the proposed DNA variants are specifically associated with TSW and, if so, what the biologic consequences of those variants might be. Similarly, the statistical association with Protopic (tacrolimus; Fig. 1A) will require targeted evaluation.

Importantly, while our case series suggests expanded clinical assessments are warranted, no patient reported complete resolution of symptoms. While prolonged treatment may yield continued improvement, preemptive caution may be warranted for exposures longer than 4 consecutive months and identifying preventive strategies and predictive factors should remain paramount.

## Data Availability

All data produced in the present study, linked to databases indicated, or are available upon reasonable request to the authors

## Acknowledgments

This work was supported by the Division of Intramural Research (DIR) of the National Institute of Allergy and Infectious Diseases, NIH. LMF and MG are supported by the Intramural Research Program of the National Institute of Arthritis and Musculoskeletal and Skin Diseases. NIAID Centralized Sequencing Program contributor information: Rajarshi Ghosh, Bryce Seifert, Mari Tokita, Jia Yan, Colleen Jodarski, Michael Kamen, Rachel Gore Moses, Nadjalisse Reynolds-Lallement, Katie Lewis, Sarah Bannon, Adrienne Borges, Nicole Gentile, Katya Damskey, Sophie Byers, Halyn Orellana, Sruthi Srinivasan. Allergy & Asthma Network has received funding from Sanofi, Regeneron, Genentech, Pfizer, and Novartis for unbranded disease awareness and education; such funding was not relevant to the presented research.

