## Supplemental Methods and Figures for "Topical Steroid Withdrawal is a Targetable Excess of Mitochondrial NAD+"

**Author Contributions**

**Supplemental Table Legends**

**Supplemental Figures**

**Supplemental References**

**Supplemental Methods**

*Survey re-analysis*

Survey data was collected and processed as previously described^1^. Statistical re-analysis was conducted in R, using the software packages: tidyverse (https://joss.theoj.org/papers/10.21105/joss.01686) and glmnet (https://www.jstatsoft.org/article/view/v033i01). Only complete surveys were included. Least absolute shrinkage and selection operator (Lasso) was used to select a subset of 154 demographic, treatment, and disease progression variables correlated with TSW development. Briefly, Lasso is similar to linear regressions but uses a penalization term that reduces the coefficients of some variables to zero. This method emphasizes variables with strong relationships to the outcome and decreases the risk of overfitting a model. During the model fitting, the penalization parameter was determined using 10-fold cross-validation.

Only participants who reported having TSW were asked about their experience with TSW symptoms. Therefore, we were not able to calculate specificity of diagnostic criteria. Instead, major criteria were selected by a clinician as symptoms which were present in at least 80% of the participants with TSW and were distinct from atopic dermatitis (AD).  Of this, we excluded the symptoms of Fatigue and Suicidal Ideation from the diagnostic criteria as these symptoms were more general.  The remaining symptoms were classified as minor criteria.  From the minor criteria we again eliminated insomnia, emotional distress, appetite changes, and reactions to new triggers because we felt that these symptoms were nonspecific, secondary to skin disease, and/or could be a manifestation of AD.  The prevalence of these symptoms across TSW participants was estimated and the optimum combination was selected, having a sensitivity of 92%.

*Patients and Controls*

Participants with TSW and heathy controls were recruited on the NIAID IRB approved clinical protocol NCT04864886. Patients were recruited in collaboration with the International Topical Steroid Awareness Network. Powering was performed using an anticipated difference in alpha diversity of the microbiome. A Chao2 was predicted as 250 in controls with a standard deviation of 25; by comparison, patients with TSW were expected to have a Chao2 of 215. For a continuous endpoint, independent sample study with an alpha error of 0.05, 22 patients per group would provide 90% power. After informed consent was obtained, participants underwent a complete history and physical. Blood work up included complete blood count with differential, chemistry panels, and IgE levels. Two 2mm skin punch biopsies were obtained. One biopsy was embedded in OCT freezing media and flash frozen using liquid nitrogen. Samples were sent to Histoserv (Germantown, MD) for sectioning on intelislides (Bruker Daltonik) and stored in a -70^O^C freezer for later use. The second biopsy was stored in RNALater (Fisher Scientific), flash frozen in liquid nitrogen, and stored at -70^O^C until ready for RNA extraction.

Slides for MALDI analysis were placed in a desiccator for 1 hr and brought up to room temperature. Matrix deposition of 2,5-dihydroxybenzoic acid matrix (Sigma Aldrich) 20 mg/mL dissolved in 70% acetone with 0.1% TFA was applied using the same robotic sprayer resulting in a matrix density of 0.003111 mg/mm^2^. Mass spectrometry was carried out in positive ion mode with an m/z range of 400 to 2,000 (high molecular weight; HMW) and then again at 20-450 m/z (low molecular weight; LMW) with a spatial resolution of 100 μm. The feature list and intensities for both LMW and HWM were combined prior to analysis.

*Cell culture*

Human follicular stem cells (CellProgen, Torrance, CA), 3T3 fibroblasts, HaCaT keratinocytes, and Schwann nerve cells (ATCC, Manassas, VA) were purchased and cultured per manufacturer’s instructions. Nicotinic acid (Sigma) in water or 10-25uM clobetasol (Sigma Aldrich) in DMSO were added to the respective culture media for select experiments. DMSO was used at a final concentration of 0.1% v/v. Cells were cultured in T25 Flacon flasks (Fisher), Seahorse plates (Agilent, Santa Clara, CA), or imbidi 8 well removable chamber slides (Fitchburg, WI). Mitochondrial stress test was performed per manufacturer instructions using Seahorse XF Pro (Agilent). For MALDI, the cells were seeded at a density of 50,000 cells/well in 8 well chamber labtek culture slides (Lab-Tek II, CC2. ThermoFisher Scientific). Three trials of the same experiment were carried out for reproducibility. Cells were washed with 1x BPS, fixed with 4% PFA for 15 minutes and washed a second time with PBS and placed in a low-pressure desiccator for 1 hour. Optical images of each well were taken prior to matrix deposition of 2,5-dihydroxybenzoic acid matrix (Sigma Aldrich) 15 mg/mL dissolved in 70% ACN with 0.1% TFA and was applied using a TM-Sprayer robotic sprayer (HTX Technologies). The estimated matrix density was 0.001611 mg/mm^2^. Mass spectrometry data was collected on a MALDI timsTOF Fleximager (Bruker Daltonik, Bremen, Germany) operated in TIMS qTOF positive ion mode from m/z 20 to 1,100 with a spatial resolution of 30 μm, mass measurement error <5 ppm, and 40,000 resolving power. All MALDI imaging data were visualized using SCiLS Lab Version 2021 (Bruker Daltonics). Denoising was carried out by moving the sliding window feature in SCiLS for the maximum number of peeks for each experiment. The average ion signals up-regulated in the experimental group vs the average signals found in the control were ordered based on threshold and important into Metaboanalyst for pathway identification using the functional analysis tool.

*Metabolic pathway analysis*

Pathway identification was performed using MetaboAnalyst (<https://www.metaboanalyst.ca/>) which determines the probability of a pathway being precent based on MS1 features associated with multiple metabolites within the reaction hierarchy of a specific pathway. The output of MetaboAnalyst (functional analysis), index of pathway significance (IPS) values was calculated as previously described^2,3^. IPS was calculated by ranking the metabolites most indicative of the TSW cohort versus both AD and HV using the ROC feature of SCiLS (Bruker); then repeated after ranking metabolites by those most predictive of the HV and AD cohort compared to TSW.

*Healthy control glucocorticoid exposure*

For controlled glucocorticoid skin exposure, healthy participants were enrolled on the NIAID IRB approved NCT02798523. 19 healthy volunteers ages 18-64 were recruited and, after informed consent was obtained, were treated with topical methylprednisolone aceponate (Advantan emulsion 0.1%; Bayer). A baseline 3mm skin punch biopsy was obtained from one arm, then topical methylprednisolone was applied to a limited area of skin in the contralateral arm. An additional 3mm skin biopsy was obtained 4 hours after drug administration, from the area where topical methylprednisolone was applied. With each biopsy, the epidermis layer was collected in tissue-grinding CKMix50-R 2ml tubes containing beads (Bertin Corp) in 500 uL TRIzol (ThermoFisher Scientific) and homogenized immediately using a Precellys 24 machine (Bertin Technologies) for 2 cycles of 5000 rpm for 20sec with a 90 sec break on ice between cycles. Samples were stored without the beads in TRIZol at -70^O^C until the time of RNA purification.

*Serum Metabolomics*

Serum was diluted 1:10 in 70% methanol (Fisher Chemical; Waltham, MA) with 0.1% trifluoroacetic acid (Thermo Fisher; Waltham, MA) and 30mcg/mL of dihydroxybenzoic acid (Sigma; St. Louis, MO). 1mcL of the matrix and serum mixture was plated on a spot plate (Bruker, Billerica, MA). Serum was run once for m/z range of 20-650, then replated and run again at 650-1700m/z. The data were analyzed separately except for analysis of ROC (performed using SCiLS; Bruker) in which the combined data set was ranked by ROC before pathway analysis by MetabAnalyst^4^. Both spot plate serum and skin biopsies were analyzed by MALDI-TOF imaging mass spec as previously described on positive ion mode^5^. Collisional cross section identification was performed using MetaboScape (Bruker). Pathway disruption was visualized using Pathview^6,7^.

*Microbiome*

Skin swabs for microbiome was performed using Floq swabs (Copan; Murrieta, CA) moistened with normal saline (Sigma). DNA was extracted and sequenced by COSMOSID (Germantown, MD) then analyzed as previously described^8^.

*Serum cytokines*

Serum cytokines were analyzed using the O-Link system (Uppsala, Sweden) performed under contract with Vanderbilt University (Nashville, TN).

*Tissue Section Preparation and Immunostaining*

Fresh frozen tissue sections of patient skin biopsy samples were fixed with BD Cytofix/Cytoperm solution (BD Bioscience, Cat#: 554722) diluted 1:4 in PBS for 30 minutes at 4°C. Healthy human skin samples were kindly provided by Dr. Jonathan M. Hernandez, Surgical Oncology Section, National Cancer Institute. Sections were washed in 1X PBS for 30 minutes before blocked in 1% BSA and 1% mouse serum in 0.1 M pH 7.4 Tris buffer containing 0.3% Triton X-100 for 30 minutes at room temperature. Then, tissue sections were incubated with directly conjugated primary antibodies (listed below) diluted in blocking solution for 10-15 h at 4°C, washed in 0.1 M pH 7.4 Tris buffer and mounted with SlowFade Gold Antifade Mounting solution (ThermoFisher Scientific Cat#S36937) using #1.5 cover glass (VWR, Cat#: 48393-241). A combination of the following fluorescent chromophores were used for immunostaining: Brilliant Violet 421, Brilliant Violet 510, Violet 450, Brilliant Violet 570, Brilliant Violet 650, PE, Alexa Fluor 488, Alexa Fluor 647, Alexa Fluor 700, Alexa Fluor 790, eFluor 570 and eFluor 615. Antibodies included from BioLegend: mouse anti-human CD1c-BV421 clone: L161, Cat# 331526; Mouse anti-human HLA-DR-BV570 clone: L243 Cat# 307638; Mouse anti-human CD69-AF700 clone: FN50 Cat# 310922; Mouse anti-human CD11c-BV510 clone:S-HCL-3 Cat# 371514. From eBioscience: Mouse anti-human FoxP3-eF570 clone: 236A/E7 Cat# 41-4777-82; Mouse anti-human CD20-eF615 clone: L26 Cat# 42-0202-82; Mouse anti-human CD103-PE clone: B-Ly7 Cat# 12-1038-42. From R&D Systems: Goat anti-human CD4-AF488 poly clone Cat# FAB8165G. From BD Biosciences: Mouse anti-human CD3-BV650 clone: SP34-2 Cat# 563916; Mouse anti-human CD8-V450 clone: RPA-T8 Cat# 560347; Mouse anti-human CD56-AF647 clone: B159 Cat#557711. From Santa Cruz Biotechnology Mouse anti-human CD68-AF790 clone: KP1 Cat# sc-20060 AF790.

*Whole Skin Preparation, Immunostaining and Clearing*

Fresh healthy skin tissue blocks were fixed in BD Cytofix/Cytoperm solution (BD Bioscience, Cat#: 554722) diluted 1:4 in PBS for 5 days at 4°C, washed in PBS 3X and left in PBS for 10 hours at 4°C. Then, the tissue blocks were transferred to PBS containing 30% glucose and left for 2 days. The tissues were then embedded in OCT and stored in -80 °C before use. A 300 micron thick skin cross sections were cut by use a Leica CM1950 cryostat. Immunostaining and clearing of the thick skin sections were performed using the protocol previously described^9^. The following fluorescent dye directly conjugated primary antibodies were used for immunostaining: CD8-V450, HLADR-AF647, CD4-AF488, CD3-iF594 and CD69-AF700.

*Confocal Microscopy Imaging*

Digital scan of both the thin and healthy thick skin sections was performed using an inverted Leica TCS SP8 X confocal system equipped with an 80 MHz pulsed white light laser, 4 Gallium-Arsenide (GaAs) Hybrid Detectors (HyDs) and 1 multialkali photomultiplier tube (PMT) with spectral detection capability. A 40X (NA 1.3) oil emersion objective lens was used for scanning the thin skin sections with a pixel size of 568.74X568.74 μm^2^ and pixel dwell time of 1.2 μs. A 20X (NA 0.75) multi-emersion objective lens was used for volume scanning of the thick skin sections with a voxel size of 1.139X1.139X2.0 μm^3^. Pixel dwell time was maintained at 1.2 μs.

*Image Processing and Visualization*

Digital images acquired by the Leica confocal system were tile stitched and then processed to correct signal spillovers from nearby channels and autofluorescence signal within the skin by using the manual unmixing method of the Automatic Dye Separation function within the Leica Application Suite X (LAS X) software package (version 4.4.0.24861). The output images (.lif files, Leica file format) were converted into Imaris version 5.5 files by Imaris software (Version 9.5.0, Bitplane) and imported into Imaris. Images of individual channels were passing through a Gaussian filter to remove random noise before pseudo-color assignment, visualization and animation creation. In all image processing steps, image size and aspect ratio were maintained identical by keeping the pixel or voxel sizes unchanged.

*IF staining TRPA1*

Human skin biopsies were freshly frozen after embedding in the OCT medium and sent out for sectioning (10µm) (Histoserv Inc., Germantown, USA). Followed by placing the slides in the desiccator to dry them and placing them into the PBS solution to remove the OCT. Skin biopsy we fixed using 4%PFA for 30 minutes, followed by washing with PBS and permeabilized with 0.5% tritone X100 for 20 minutes. Blocking was performed using with 5% Normal Goat Serum (NGS) for 1 hour. Primary antibody (#ACC-037 Alomone labs) solution was added according to the manufacturer’s instruction at 4°C overnight. Slides were washed 3 times for 15 minutes each in PBS. A secondary antibody (Alexa flour 488 or 568) solution was added according to the manufacturer's instructions for 1 hour at room temperature. Slides were washed 3 times for 15 minutes each in PBS. DAPI solution was added and incubated for 15 minutes at room temperature. Slides were washed 3 times with PBS and were mounted with mounting medium.

*Genome sequencing*

Genome sequencing and analysis was performed as previously described^10^. VCF files were filtered using bcftools^11^. CADD scores were calculated for all variants^12^. Genes for which at least 12 of 16 patients had at least one heterozygous or homozygous variant with a PHRED-scaled CADD score of 20 were submitted to STRING for functional enrichment analysis^13^. These genes were also cross-referenced to RNA-seq data to identify variants which were associated with changes in gene expression. Gene maps of the identified genes, *PAX6* and *FMO2*, as well as *TTN* were generated using trackViewer in R^14^.

*RNAseq*

For biopsies of healthy controls exposed to glucocorticoids in a controlled setting, punch biopsies were homogenized for 40s in lysing matrix D tubes (MP Biomedicals, Santa Ana, CA) containing 1,000μl Trizol (Thermofisher Scientific, Waltham, MA) in a FastPrep® FP 120 instrument (MP Biomedicals) at speed 6.0 meters per second. Homogenized Trizol lysate was combined 1-Bromo-3-chloropropane (MilliporeSigma, St. Louis, MO), mixed, and centrifuged at 16,000 x *g* for 15 min at 4°C.  RNA containing aqueous phase was collected from each sample and passed through QIAshredder column (Qiagen, Valencia, CA) at 21,000 x g for 2 minutes to homogenize any remaining genomic DNA in the aqueous phase.  Aqueous phase was combined with equal amount of RLT lysis buffer (Qiagen, Valencia, CA) with 1% beta mercaptoethanol (MilliporeSigma, St. Louis, MO) and RNA was extracted using Qiagen AllPrep DNA/RNA mini columns (Valencia, CA). RNA integrity was assessed using the Agilent 2100 Bioanalyzer using RNA 6000 Pico kit (Agilent Technologies, Santa Clara, CA). RNA was quantitated using a fluorescence assay (Quant-it RiboGreen RNA, Thermofisher Scientific, Waltham, MA) on a Tecan Spark multiplate reader (Tecan, Switzerland).

Sequencing libraries were generated using the SMARTer Stranded Total RNA-Seq Kit v3- Pico Input Mammalian (Takara Bio USA, Inc., San Jose, CA) following the manufacturer’s protocol without modification. RNA input for NGS library preparation was 10 ng for the TSW samples and 3.75 ng for the punch biopsy samples. The TSW samples were amplified for 12 cycles and the punch biopsy samples were amplified for 13 given the amount of RNA in the preparation. Microfluidic electrophoresis for library quality control was performed on a TapeStation 4200 (Agilent Technologies, Santa Clara, CA) with D1000 screen tapes. Two microliters of each library were combined and sequenced on the MiSeq (Illumina, San Diego, CA) using a Micro v2 300 cycle chemistry kit to acquire the reads/uL for each library. The MiSeq data was used to create a normalized library pool for sequencing on the NovaSeq X Plus (Illumina, San Diego, CA) with 200 cycle chemistry.

Using the CogentAP NGS Analysis v2.0 pipeline (<https://www.takarabio.com/products/next-generation-sequencing/bioinformatics-tools/cogent-ngs-analysis-pipeline> ), we first processed raw fastq files to trim UMIs and append UMI sequence to the read ID. The pre-processed fastq files were then trimmed for quality and adapter contamination using Cutadapt v4.0^15^. Trimmed reads were mapped to the hg38 reference genome and Gencode GRCh38 v.42 transcriptome using STAR v2.7.6a^16^ in two-pass mode, and PCR duplicates were flagged using Picard MarkDuplicates from GATK4 v4.2.6.0^17^. Gene-level expression quantification was performed using RSEM v.1.3.0^18^. Differential gene expression was carried by using DESeq2 package in R. Functional enrichment analysis of significantly different transcripts was performed using STRING. Pathway disruption was visualized using Pathview.

*Mice*

Male and female BALBc/J mice aged 6-11 weeks (age and sex matched within each experiment) were purchased from Jackson Labs (Bar Harbor, MN). Nicotinic acid (Sigma) was diluted in water to final concentrations of 5M, 500mM, and 5mM and applied topically to each mouse ear at 10mcL per daily for 8-10 days. Thickness was measured using calipers (Matsui; Shiga, Japan) on day 9-11.

*Berberine dilution*

1 capsule of berberine (Solaray; Jim Beck, Utah) was dissolved in 1L of sterile water (Sigma). Based on the molecular weight, the resultant concentration was calculated at 1.5M. Dilutions were performed to achieve 0.6uM to align with the cell culture lowest effective dose. Images were taken after 15min of dilution.

**Data availability:**

Shotgun metagenomics and RNA seq datasets generated during the current study are available in the NCBI under the BioProject ID: PRJNA1098044. Metabolomic data will be accessible via MetaboLights accession number MTBL9913.

**Author contributions**:

NS clinically evaluated the patients and wrote the manuscript, SS wrote the manuscript, GR generated the graphs for the survey results, variant analysis, and metabolomics, MY performed immune-staining and MALDI mass spec, PPC analyzed shotgun metagenomics, RNAseq and metabolomic data, AAS coordinated the patient’s visits, MG, LMF, and KNH performed the healthy volunteer glucocorticoid exposure study, SG assisted with immune-stain imaging for TRPA1, KJM performed Seahorse experiments, APH provided technical guidance on genome assessment, KK and SR performed RNA extraction and library prep, JL analyzed RNAseq data, WY performed the immune-staining for cell type, MS and MAW sequenced the patient genomes, DG, KB, and KT conducted the patient survey, and IAM oversaw the project, assessed the patients and wrote the manuscript.

**Supplemental Table Legends:**

**Supplemental Table 1:** Demographics of patients enrolled in study including treatments opted for and results. Age, gender, and ethnicity removed per medRXiv policies; the final publication will report these data.

**Supplemental Table 2:** Full output from O-LINK assessment of serum cytokines in patients with topical steroid withdrawal (TSW), atopic dermatitis without TSW (AD), and healthy controls (HV).

**Supplemental Table 3:** Identified variants in mitochondrial genes for TSW cohort.

**Supplemental Figures**

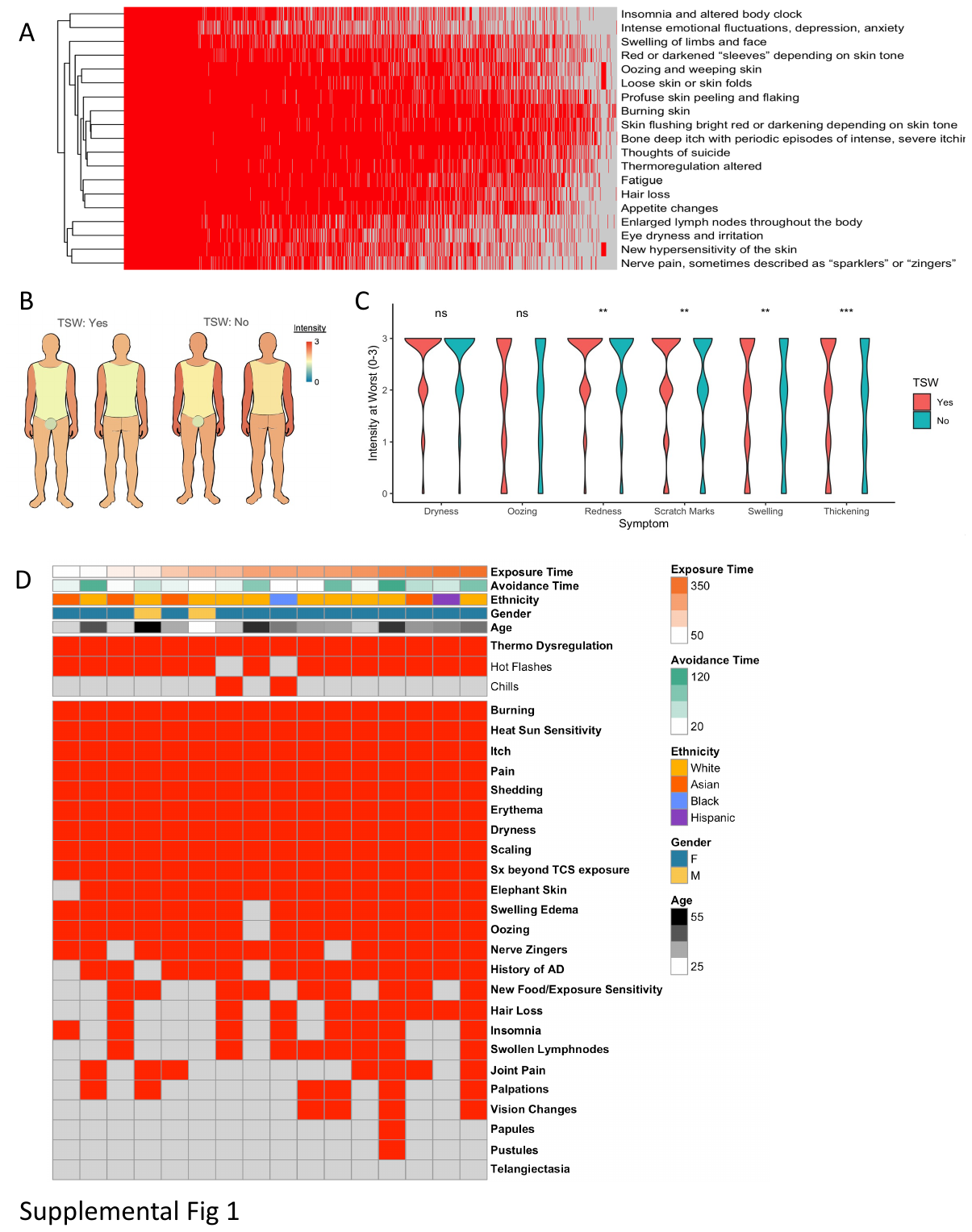

**Supplemental Figure 1: Topical Steroid Withdrawal (TSW) is distinguishable from atopic dermatitis (AD).** (A) Full responses from symptoms asked in prior survey of 1,486 patients with eczematous phenotype reporting TSW. (B-C) Intensity by body site (B) and self-reported PO-SCORAD parameter values (C) by survey from 1,889 respondents with eczematous skin disease for those reporting TSW (N=1,486) from those that denied the diagnosis. (C) Results from our cohort (N=16) indicating history of reported symptoms (red = yes, gray = no) as well as demographics for total time exposed to topical steroids, time since peak symptoms and enrollment (avoidance time), self-reported ethnicity, gender, and age.

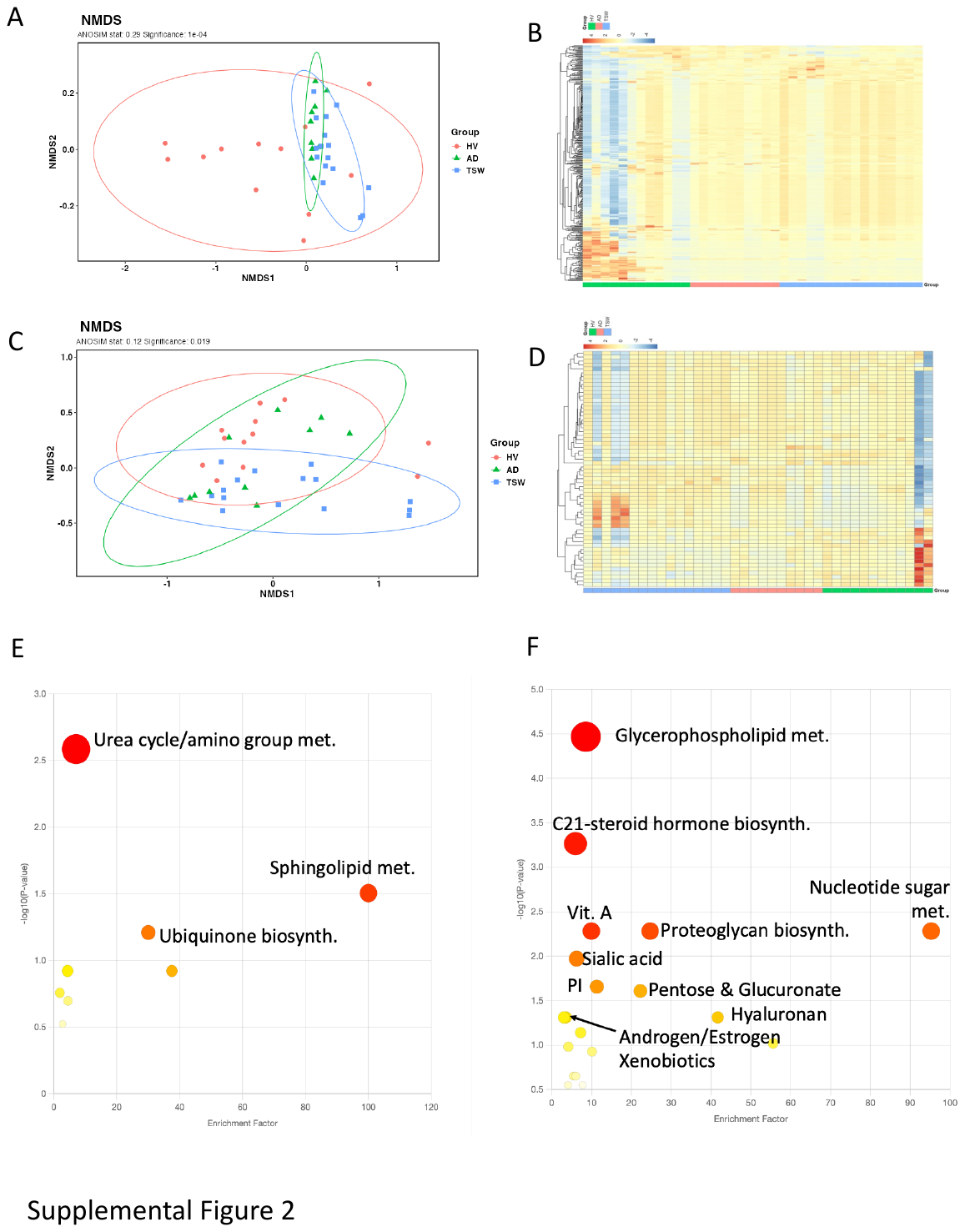

**Supplemental Figure 2: Serum metabolomics in TSW differs from AD.** (A-D) Similarity plot as measured by NMDS (non-metric multidimensional scaling; A, C), intensity heat maps (B, D) for metabolites identified by m/z and collisional cross section, and MetaboAnalyst pathway analysis via ranking metabolite profiles by differences from TSW (E-F) for serum on low molecular weight (20-450m/z; A-B, E) and high molecular weight (400-2000m/z; C-D, F) range. Comparing patients with TSW (N=16), atopic dermatitis with no history of TSW (N=10) and healthy controls (HV; N=11).

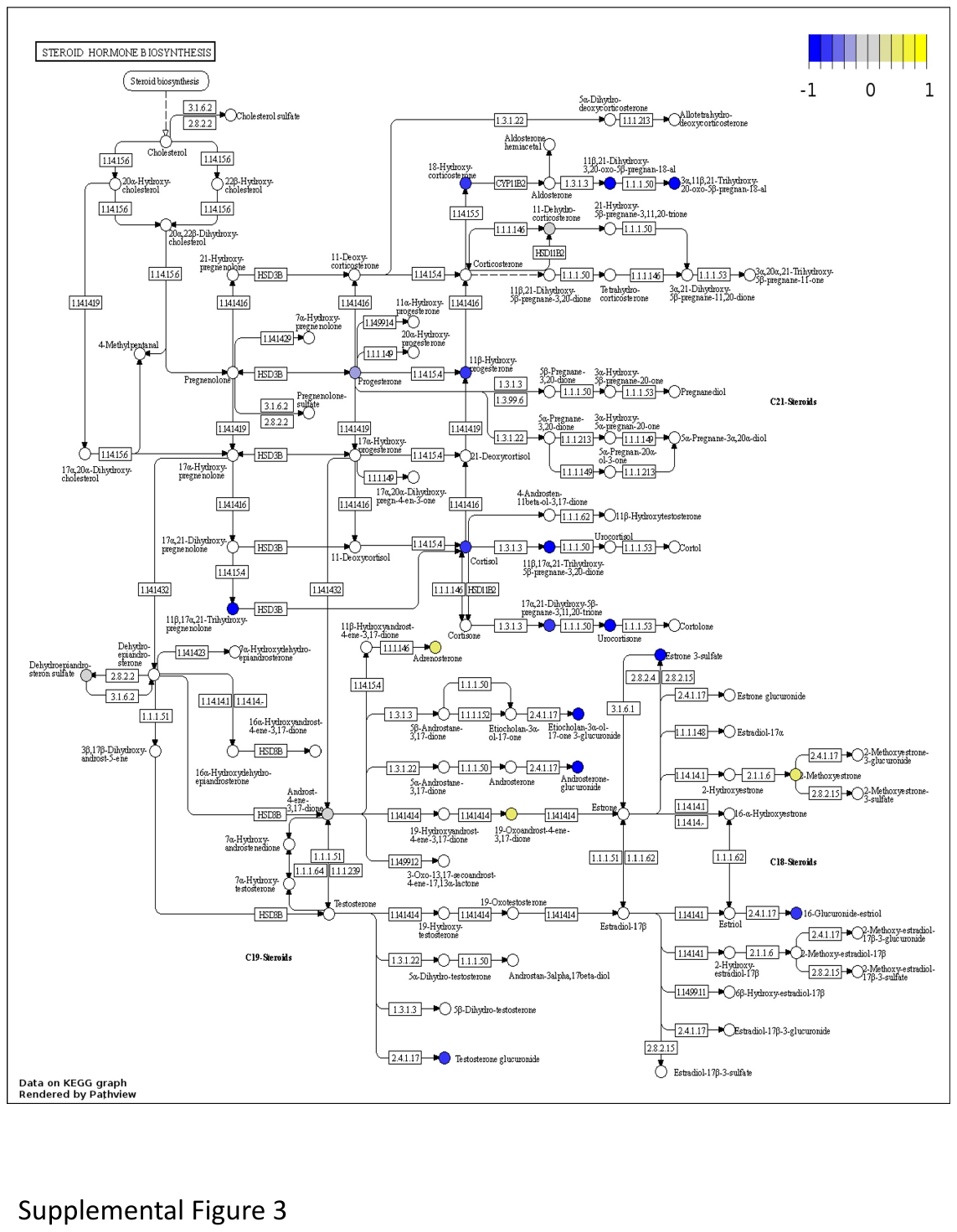

**Supplemental Figure 3: Patients with TSW have disruptions in Steroid Hormone Biosynthesis.** Steroid hormone biosynthesis pathway from KEGG with superimposed log_10_ fold change in intensity values for indicated metabolites for TSW (N=16) versus HV (N=11) cohorts.

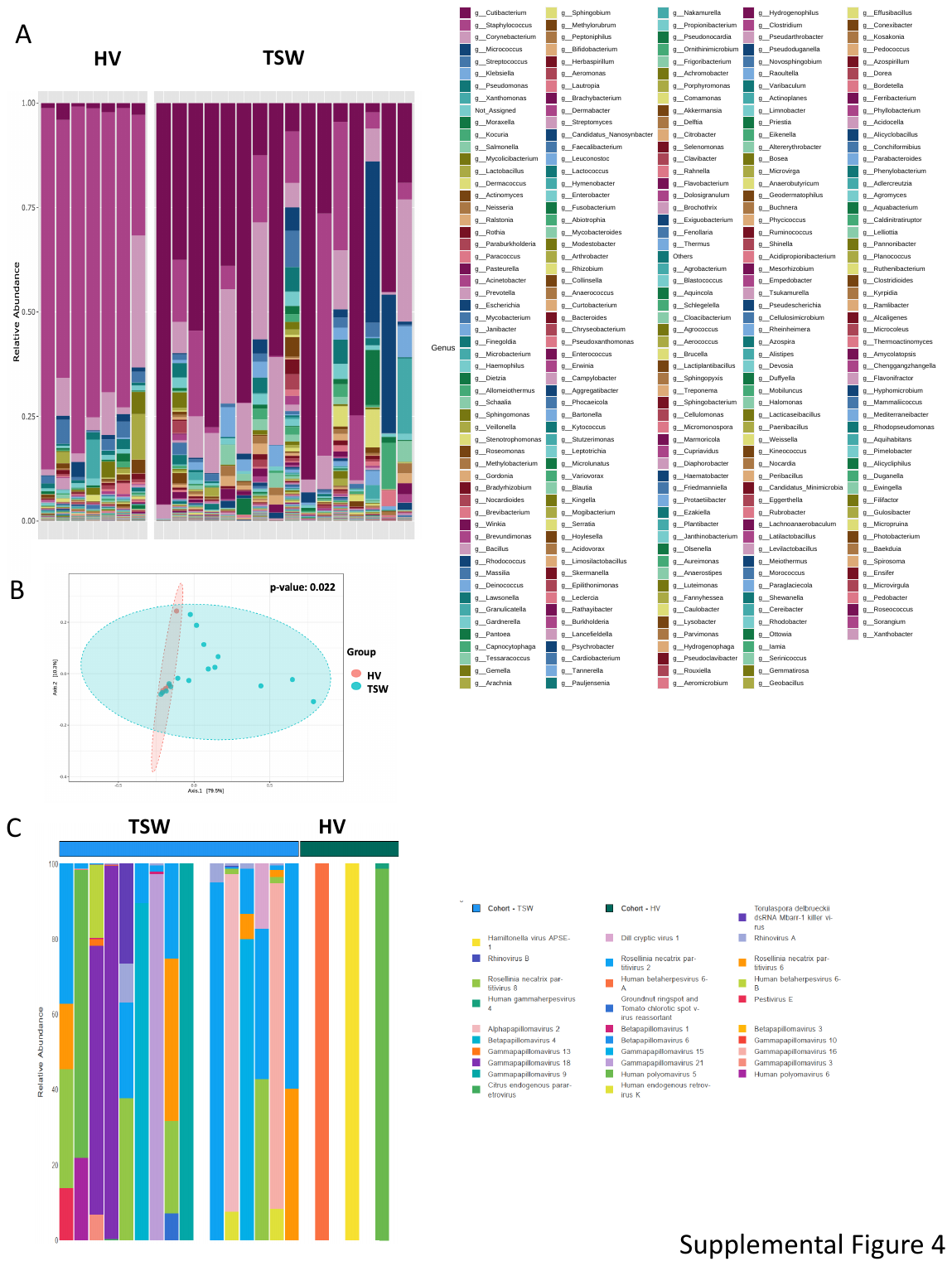

**Supplemental Figure 4: Microbiome signatures in TSW differ from healthy controls.** Shotgun metagenomic sequencing was performed on skin swabs from patients with TSW (N=16) versus healthy volunteers (N=7). (A) Full speciation at genre level as presented in Figure 2A. (B) Similarity plot as measured by NMDS (non-metric multidimensional scaling) for genre-level fungal analysis. (C) Speciation for viral microbes detected.

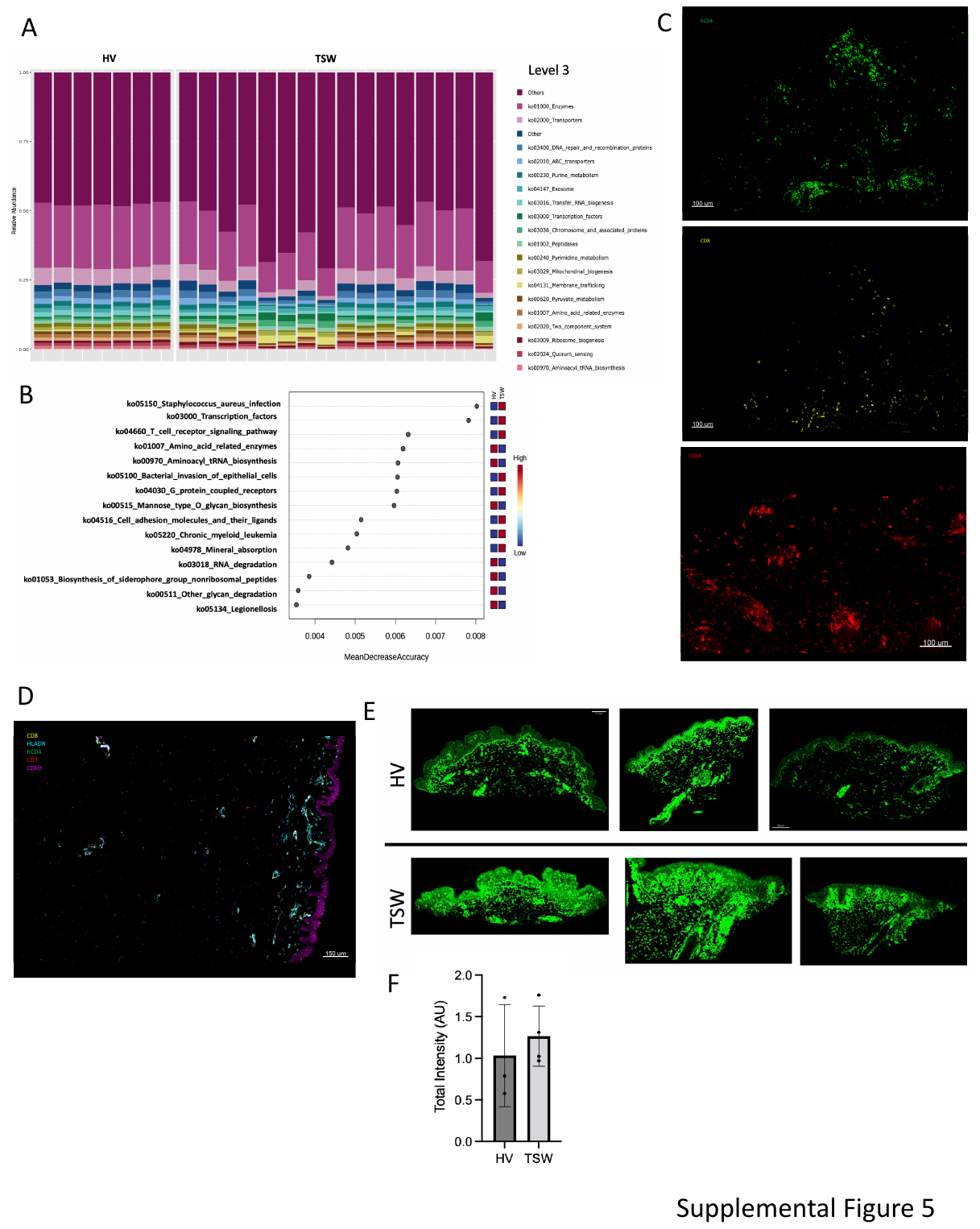

**Supplemental Figure 5: Cell infiltrates in TSW are similar to reported AD findings.** Shotgun metagenomic sequencing was performed on skin swabs from patients with TSW (N=16) versus healthy volunteers (N=7). (A) KEGG level 3 analysis for functional analysis on full microbial DNA (B) Random Forest plot for predictive functional genes for TSW versus healthy control (HV). High in the indicated group is depicted in red boxes, lower values in the indicated group is depicted by blue boxes. (C) Representative images for stains for CD8, human CD4, and CD68 in patient with TSW. (D) Similar to C but for a representative HV. T cells are hardly visible in a 2D optical section view of skin cross section but are abundant when using 3D volume imaging to enhance cell detection (See Supplemental Video 1). (E-F) Stains (E) and total intensity values ± SD (F) for 3 patients with TSW and 3 HV for the ion challenge TRPA1.

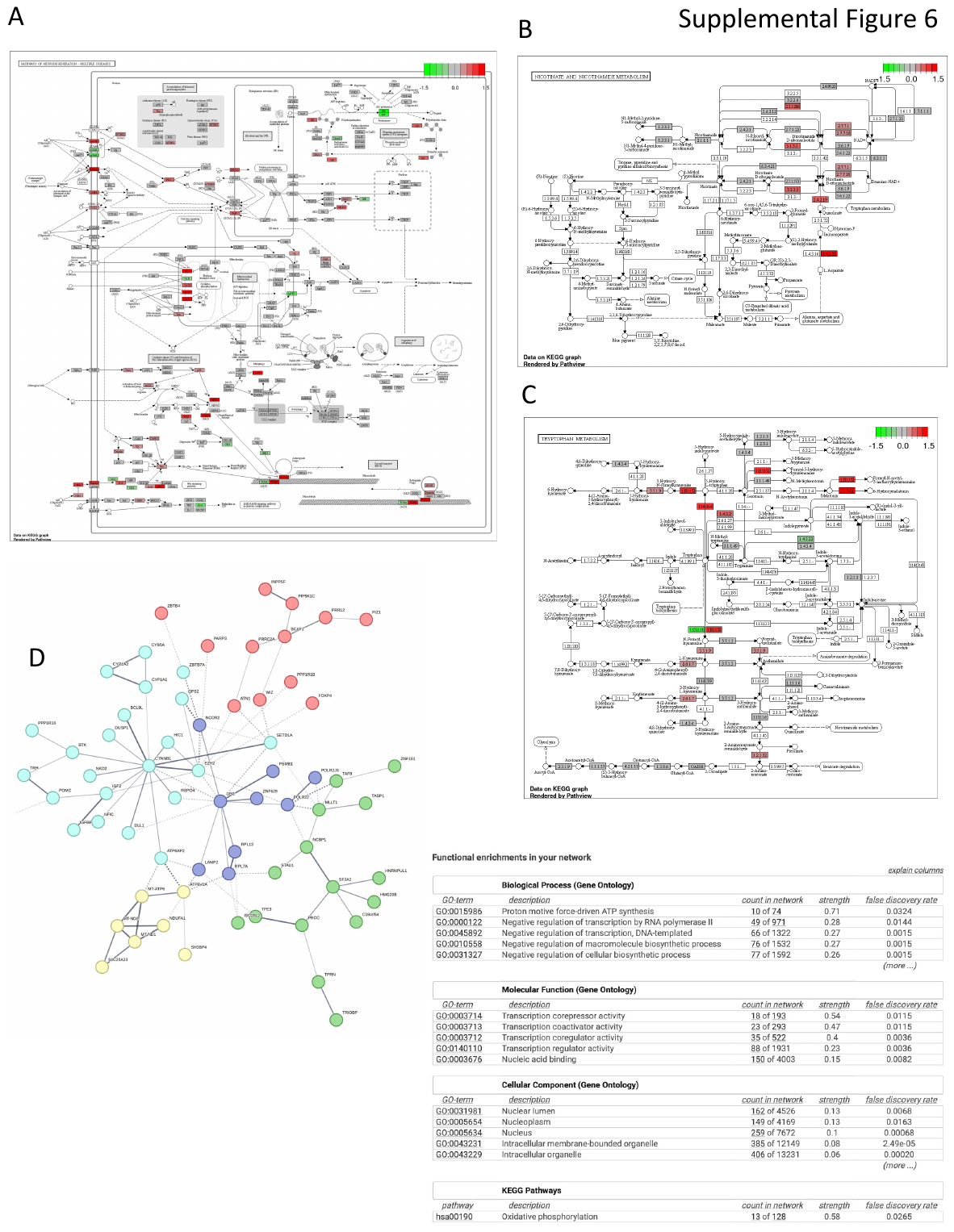

**Supplemental Figure 6: Skin from patients with TSW demonstrates disruptions in Niacin related metabolic pathways.** Skin biopsy samples for patients with topical steroid withdrawal (TSW; N=16) and healthy volunteers (HV; N=11) underwent RNAseq analysis. (A-C) KEGG pathway analysis for Neurodegeneration (A), Nicotinate and Nicotinamide (B), and tryptophan metabolism (C). Superimposed log_10_ fold change, green indicates product is more abundant in HV, red indicates product is more abundant in patients with TSW. (D) STRING analysis output for most differentiating transcripts by RNAseq along with functional network estimates. Coloring indicates k means clusters provided by STRING to make five groupings.

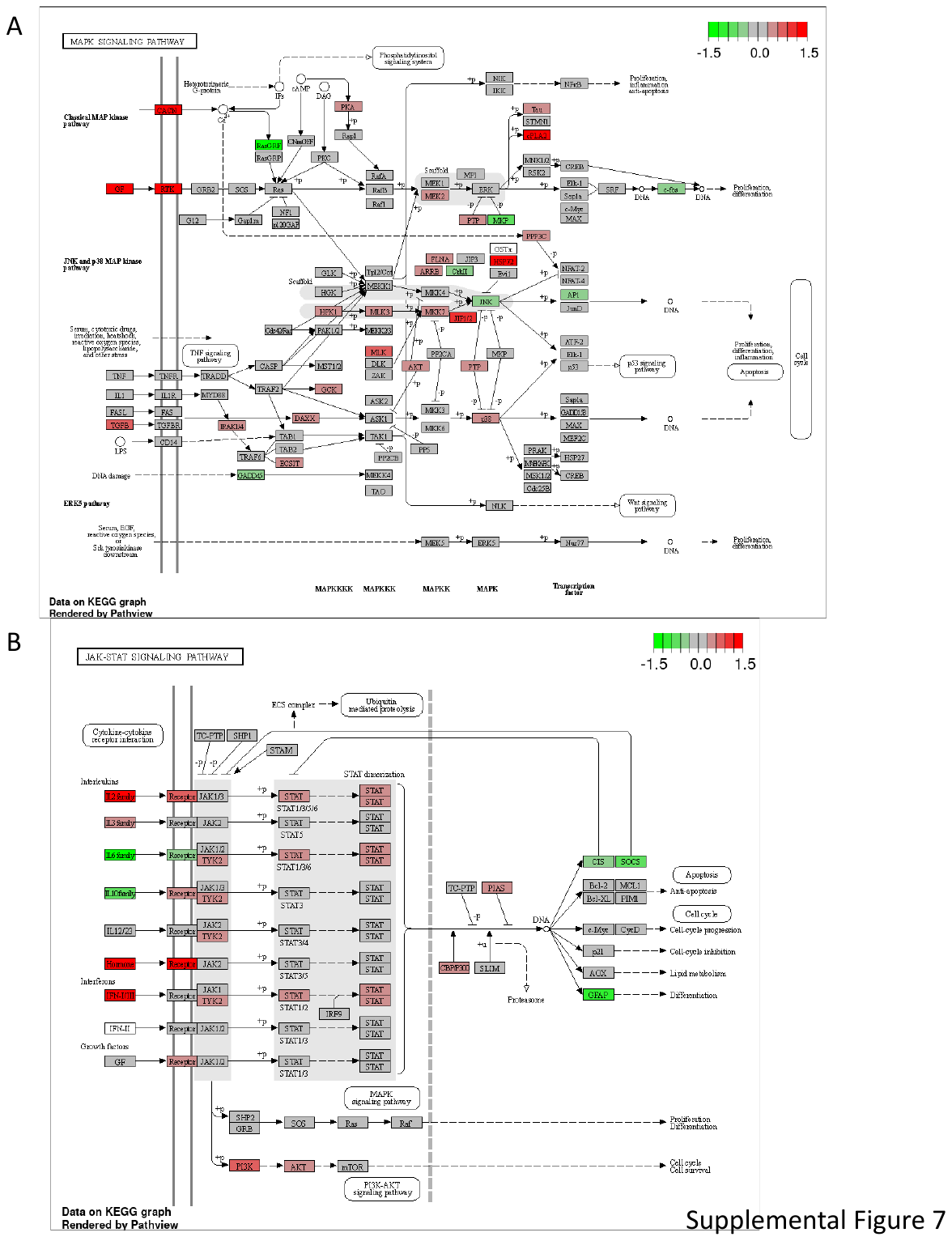

**Supplemental Figure 7: Skin from patients with TSW demonstrates disruptions in inflammatory pathways.** Skin biopsy samples for patients with topical steroid withdrawal (TSW; N=16) and healthy volunteers (HV; N=11) underwent RNAseq analysis. (A-B) KEGG pathway analysis for MAPK signaling (A) and JAK-STAT pathway (B). Superimposed log_10_ fold change, green indicates product is more abundant in HV, red indicates product is more abundant in patients with TSW.

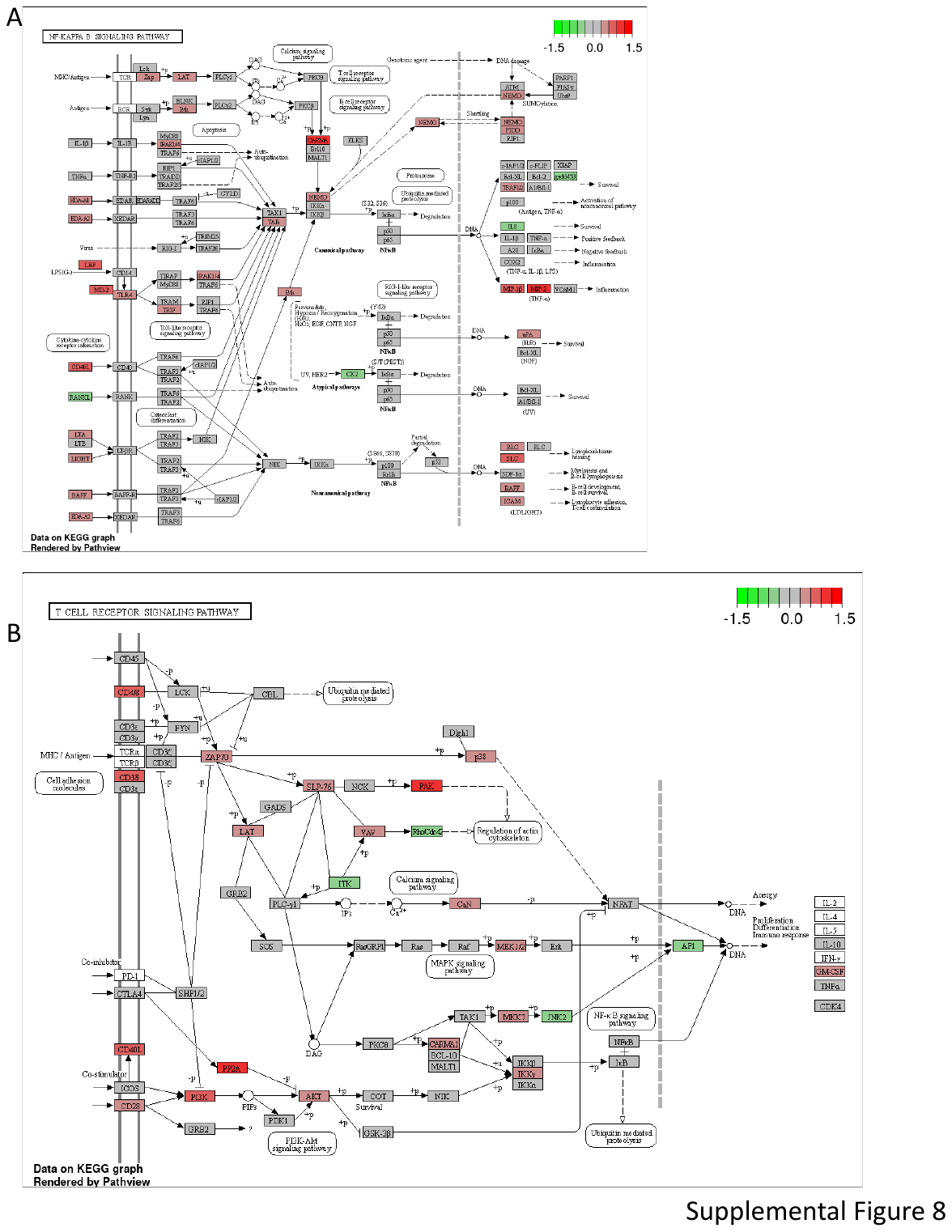

**Supplemental Figure 8: Skin from patients with TSW demonstrates disruptions in additional inflammatory pathways.** Skin biopsy samples for patients with topical steroid withdrawal (TSW; N=16) and healthy volunteers (HV; N=11) underwent RNAseq analysis. (A-B) KEGG pathway analysis for NFkB signaling (A) and T-cell receptor signaling pathway (B). Superimposed log_10_ fold change, green indicates product is more abundant in HV, red indicates product is more abundant in patients with TSW.

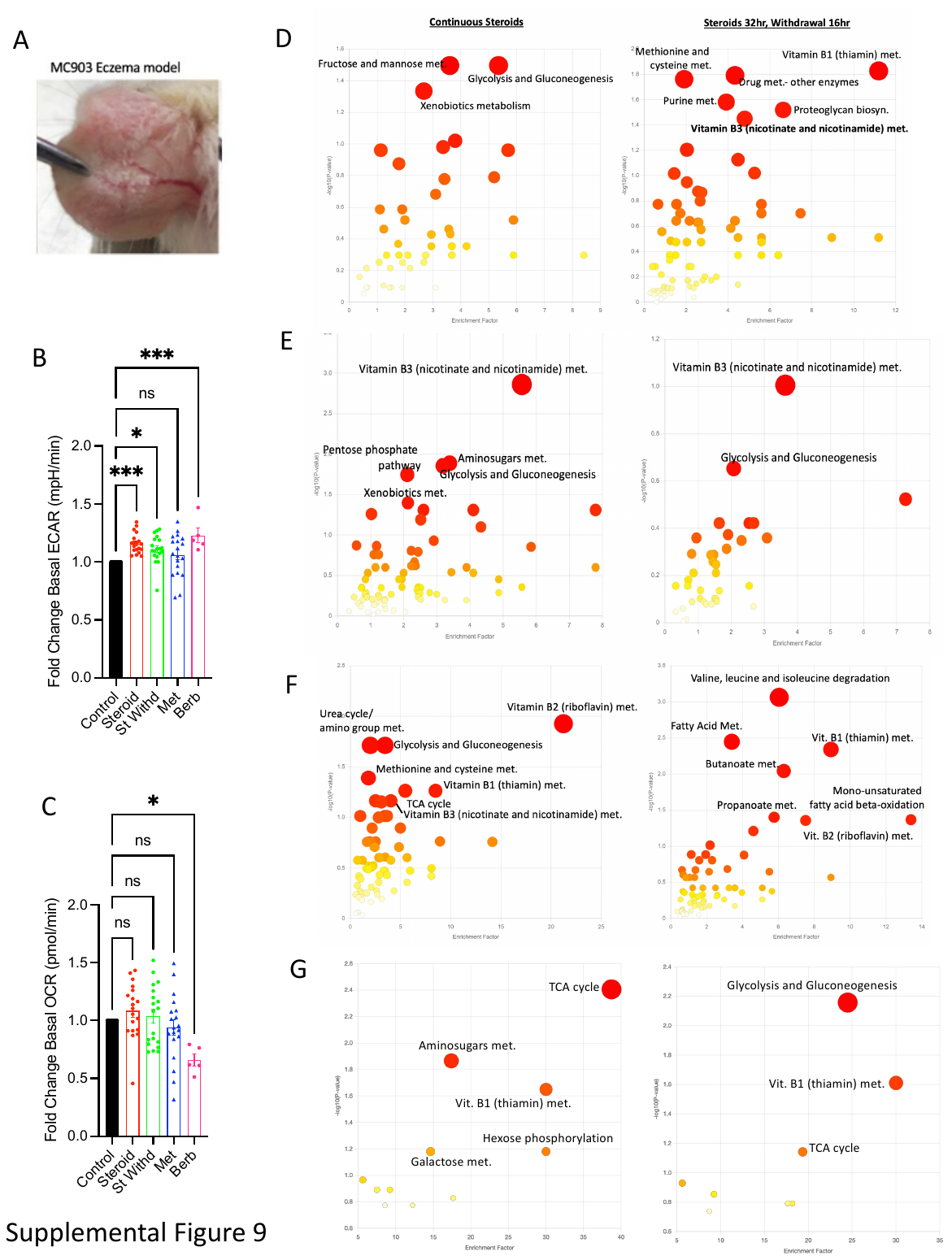

**Supplemental Figure 9: Steroid withdrawal induces mitochondrial associated niacin overproduction in human skin cells**. (A) Representative image of mouse ears under MC903 model of atopic dermatitis. (B-C) Fold change in basal extracellular acidification rate (ECAR; a measurement of glycolysis; B) and basal oxygen consumption rate (OCR; a proxy for oxidative phosphorylation; C) for Human follicular stem cells (HFSC) incubated in clobetasol for 48 hours (Steroid), clobetasol for 24 hours followed by normal media for 24 hours (Steroid withdrawal), or under steroid conditions with 10uM for metformin or berberine. Dots indicate replicate wells. (D-G) Raw MetaboAnalyst output for HFSC (D), keratinocytes (E), Fibroblasts (F), or Schwann nerve cells (G) under steroid and steroid withdrawal conditions. Data represent 3 independent experiments (A-I) and are indicated by mean ± SEM. * = p value <0.05, 888 p <0.01, ns = not significant, other p values indicated.

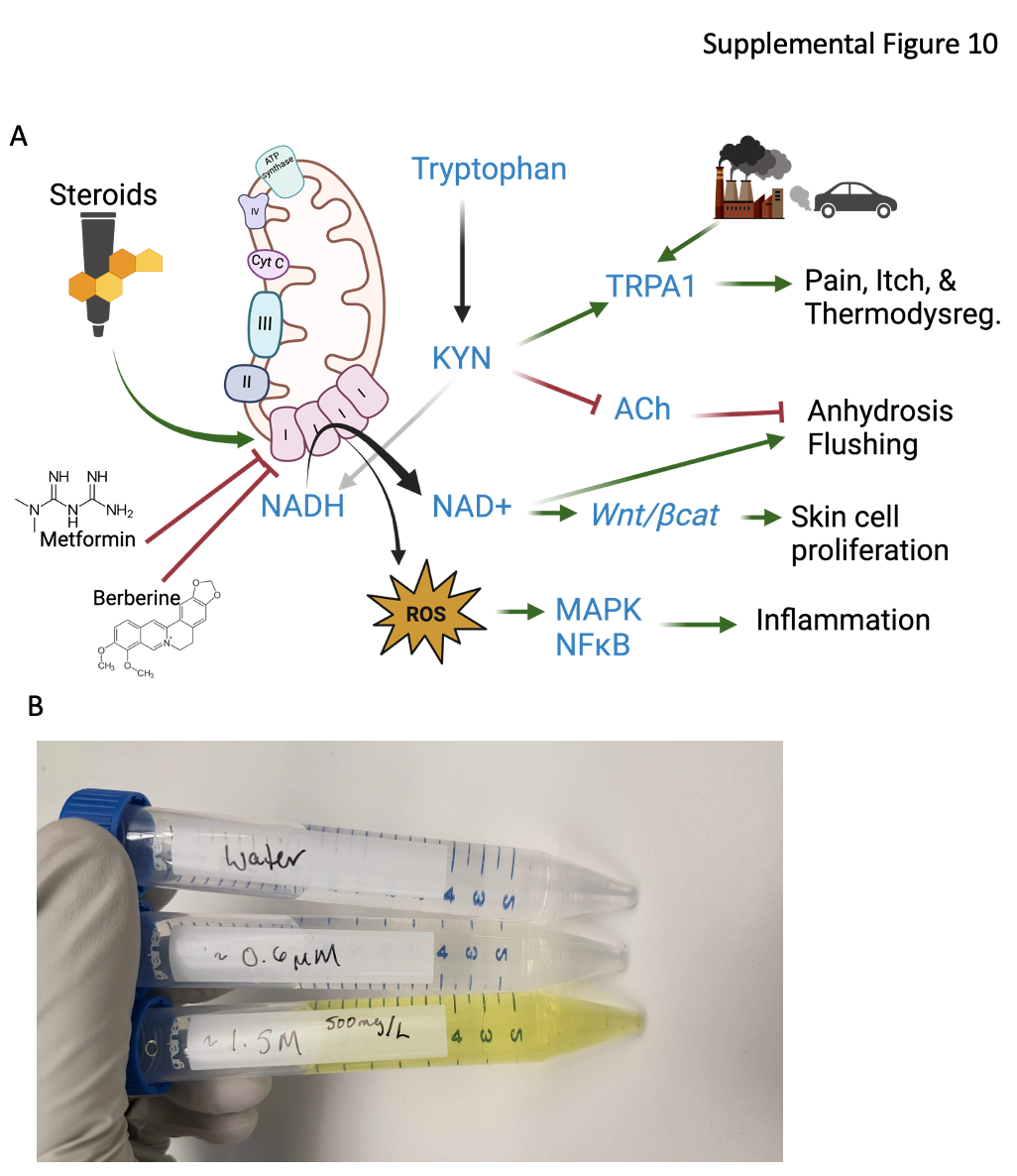

**Supplemental Figure 10: Diagram of proposed mechanism of TSW.** (A) Diagram of hypothesized mechanism of TSW pathology, image generated using BioRender.com. (B) Picture of coloration of berberine in water at indicated concentrations.

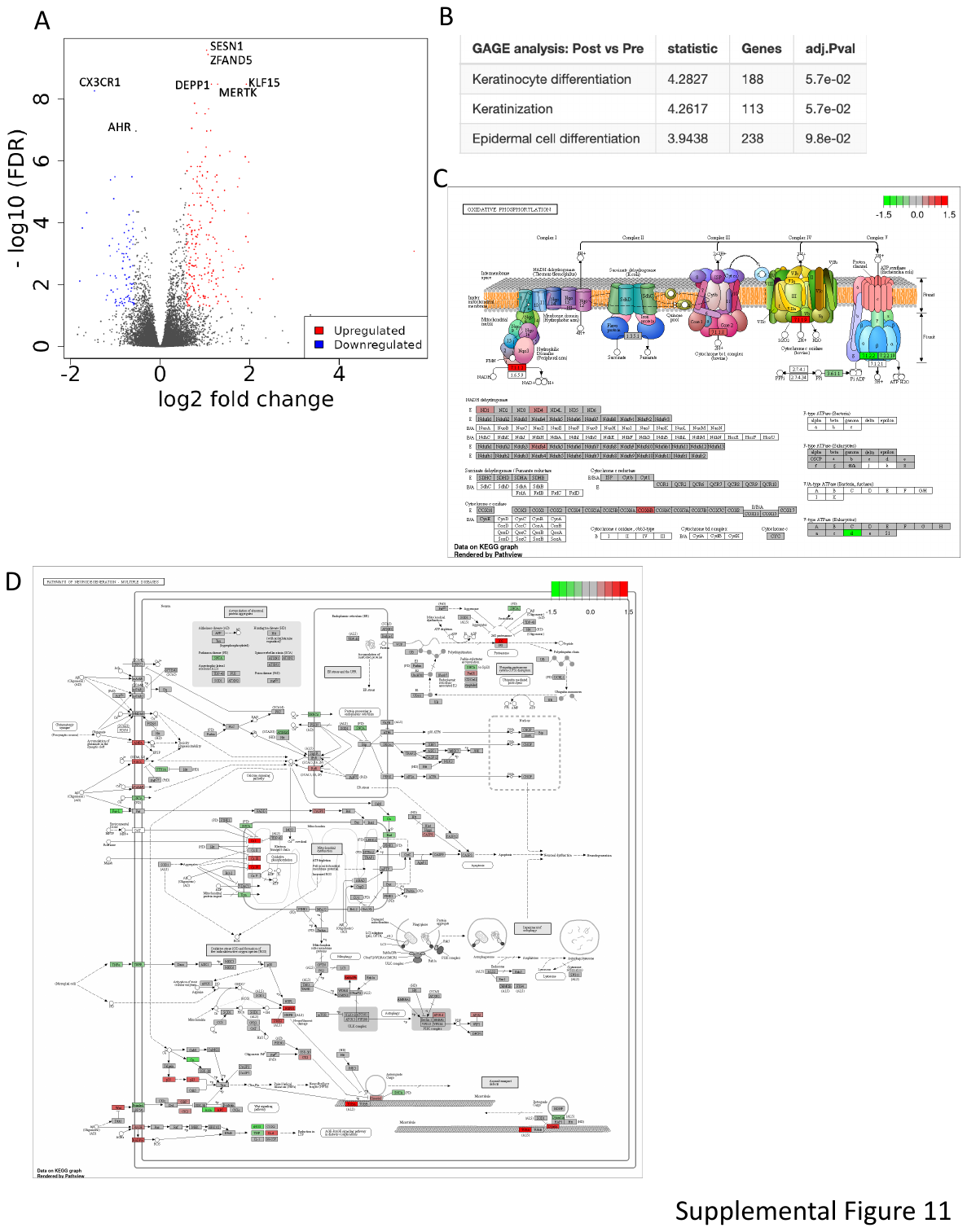

**Supplemental Figure 11: Short term exposure to topical glucocorticoids induces mitochondrial complex I**. 19 healthy controls underwent baseline skin biopsy followed by topical exposure to a glucocorticoid (see methods). 4 hours later, a repeat biopsy was performed in the area of glucocorticoid exposure. RNA-seq was performed, followed by differential expression analysis contrasting post- and pre-treatment skin transcriptomes. Volcano plot (A) and derived pathways (B) are shown. (C-D) KEGG pathway for oxidative phosphorylation (C) and neurodegeneration (D) with superimposed log_10_ fold change. Green indicates product is more abundant before glucocorticoid exposure and red indicates product is more abundant after glucocorticoid exposure.

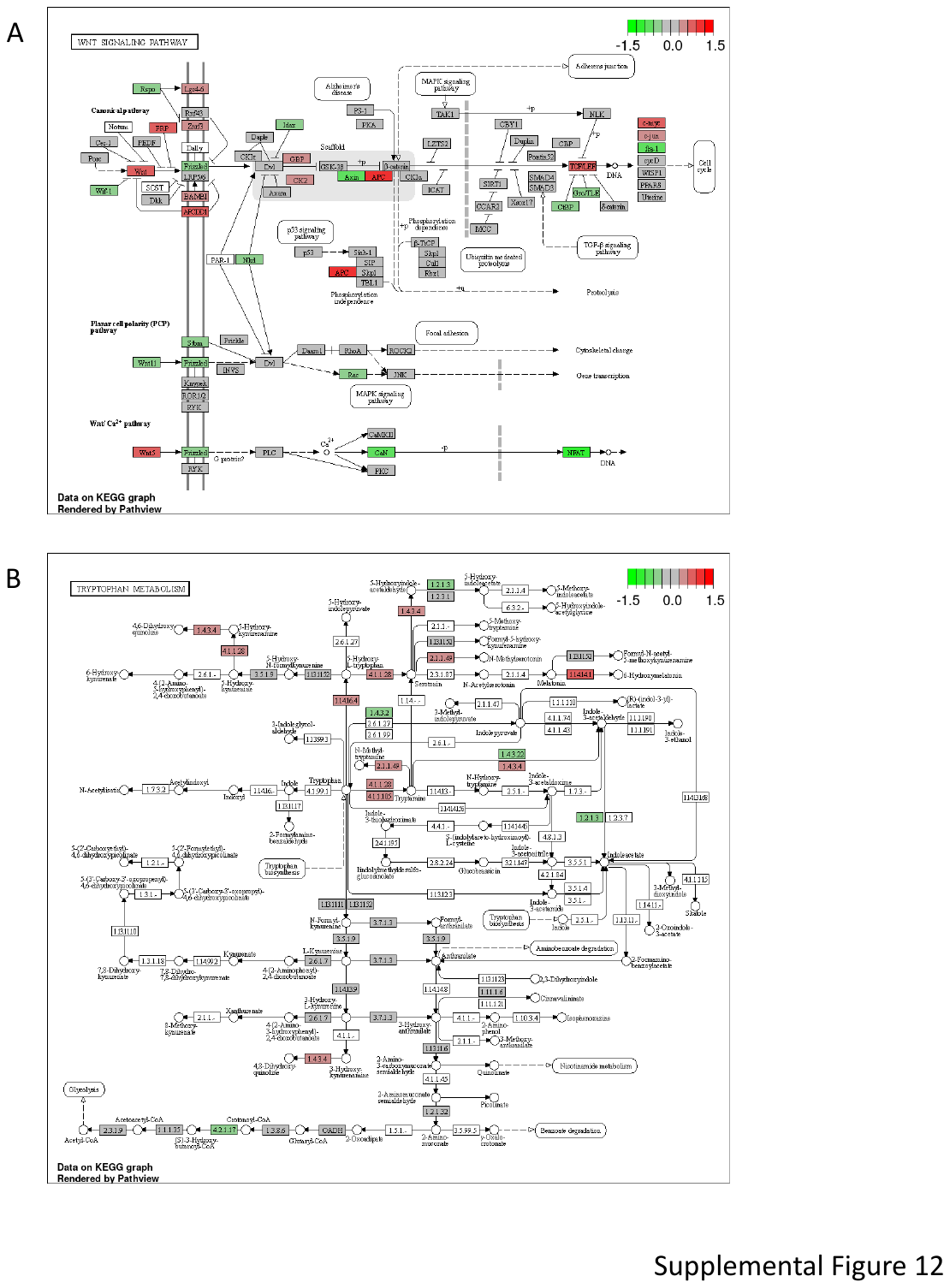

**Supplemental Figure 12: Short term exposure to topical glucocorticoids alters *Wnt* and tryptophan metabolism**. 19 healthy controls underwent baseline skin biopsy followed by topical exposure to steroids (see methods). 4 hours later, a repeat biopsy was performed in area of steroid exposure. RNAseq analysis indicated differentially expression for transcripts upregulated or downregulated by steroid exposure. (A-B) KEGG pathway for *Wnt* signaling (A) and tryptophan metabolism (B) with superimposed log_10_ fold change, green indicates product is more abundant before steroid exposure, red indicates product is more abundant after steroid exposure.

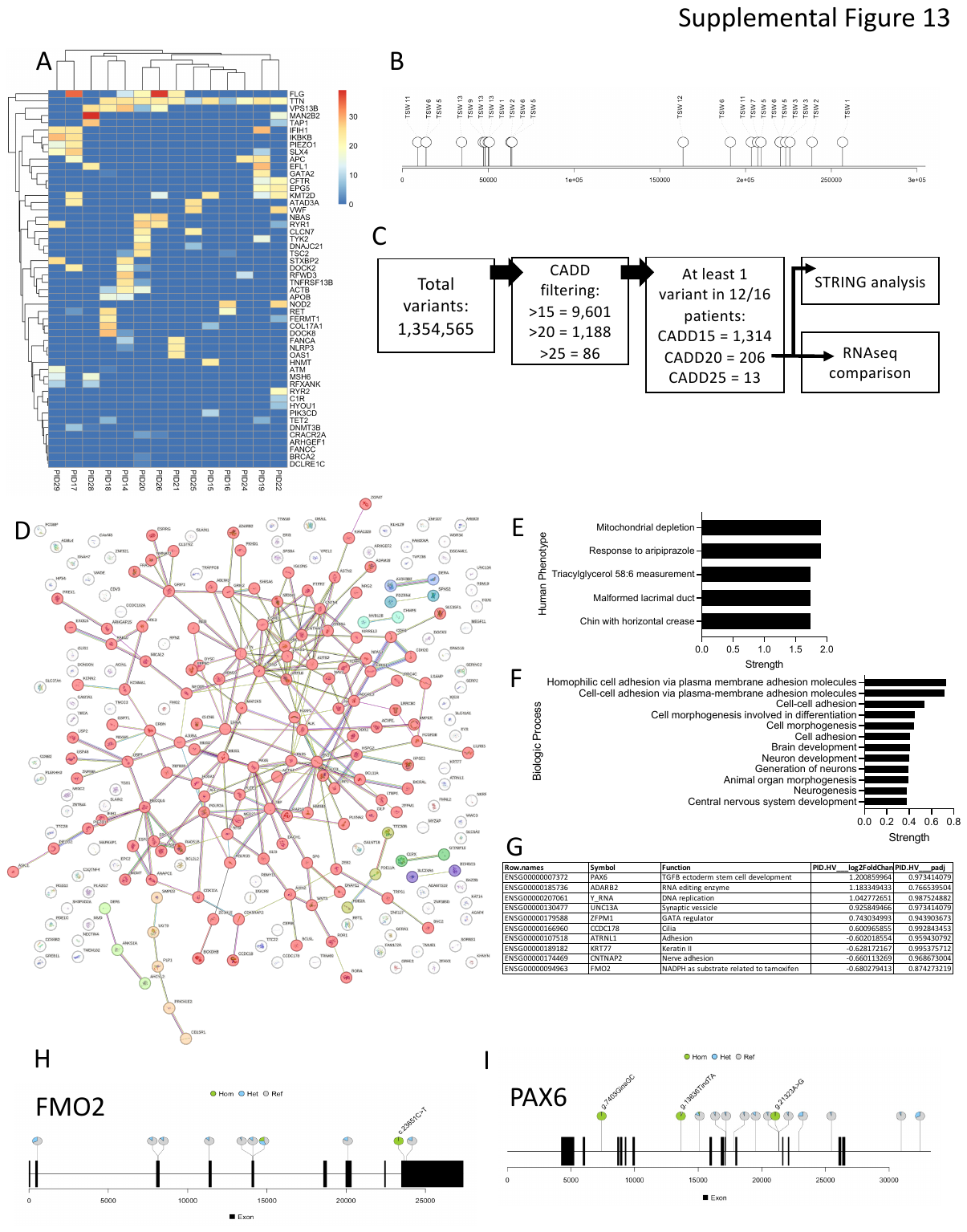

**Supplemental Figure 13: TSW associated with genetic variants in mitochondrial, neuron, and cell adhesion pathways.** (A) For the 16 patients in the TSW cohort, CADD score values for variant of uncertain significance (VUS) are shown. (B) Gene map for *TTN* for location of variants of any CADD score. (C) schema of analysis of full variants for entire cohort. (D-F) STRING analysis of 206 genes containing DNA variants with CADD score above 20 that were present in at least 12 of 16 participants. Connection plot (D), reported phenotype (E), and biologic process (F) are shown. Coloring indicate k means squared groupings by STRING into 5 groups. (G) Variants with CADD scores over 20, ranked for largest differential expression on RNAseq analysis. (H-I) Gene maps for *FMO2* and *PAX6*.
